# Causal discovery identifies posttraumatic stress as a driver of internalizing symptoms across independent veteran and civilian populations

**DOI:** 10.1101/2020.10.19.20186981

**Authors:** Benjamin Pierce, Thomas Kirsh, Adam R. Ferguson, Thomas C. Neylan, Sisi Ma, Erich Kummerfeld, Beth E. Cohen, Jessica L. Nielson

## Abstract

**Background:** Approximately half of patients with posttraumatic stress disorder (PTSD) also meet criteria for internalizing disorders, yet few studies assess reciprocal longitudinal relations among these symptoms.

**Methods:** We used longitudinal causal discovery in a veteran cohort for hypothesis-generation about PTSD and internalizing symptom drivers (n=240), followed by hypothesis-testing in two independent civilian cohorts with similar symptom assessments over time (n=79 and n=116).

**Results:** In the veteran cohort, causal discovery revealed PTSD symptoms drove internalizing symptoms, which subsequently impacted social functioning; all independent of problematic alcohol use. This replicated in treatment-seeking anxiety disorders (AD, n = 79) and substance abuse (SA, n = 116) samples with significantly better model fit for PTSD symptoms driving internalizing symptoms, versus internalizing symptoms driving PTSD symptoms (BIC change for AD sample = 175.1, p<.001; BIC change for SA sample = 571.6, p<.001). We also found better model fit with PTSD symptoms driving anxiety symptoms, versus anxiety symptoms driving PTSD symptoms (BIC change for AD sample = 71.8, p < .001; BIC change for SA sample = 568.9, p < .001). Posthoc analysis in the veteran sample revealed that hyperarousal and cognitive and affective disturbance bridged between other PTSD symptoms and internalizing symptoms.

**Conclusions:** Our findings suggest that internalizing symptoms that emerge in the context of PTSD are more likely to be driven by PTSD symptoms. These results highlight the need for a PTSD- and trauma-informed approach to treating internalizing symptoms, and provide preliminary evidence for cognition and mood disruption as a factor driving comorbidity.

## INTRODUCTION

Posttraumatic stress disorder (PTSD) is highly co-morbid with internalizing conditions, including major depression and anxiety disorders. Prevalence estimates suggest over half of PTSD patients meet criteria for one such disorder(1–8). Patients with comorbid PTSD and depression have more suicide attempts and a twofold increase in medical costs, compared with either disorder alone (9–16). Similarly, comorbid anxiety disorders are associated with more severe PTSD and impairment(7,9), often emerging in triple comorbidity with PTSD and depression(9,17). There is debate about interpreting comorbidity among PTSD and other internalizing disorders(18), including whether PTSD symptoms drive internalizing symptoms, or vice versa, or if the relationship is bidirectional (19). Studying the connections between these symptom domains and their impacts on patients’ health and functioning can support more effective, targeted care.

Elucidating the relationship between PTSD and other internalizing conditions has been challenging and hampered by attempts to isolate symptoms of each disorder (20). Many clinical studies of PTSD exclude patients with internalizing comorbidities, or treat such symptoms as confounded, while studies of internalizing disorders often exclude patients with PTSD, or neglect to evaluate traumatic history (20). Cross-sectional studies have identified shared features of PTSD and other internalizing concerns (21–25), including network analyses using an adult cohort with depressive symptoms and trauma history, which found that impaired concentration, sleep problems, irritability, and guilt, were shared across disorders (26). Other cross-sectional analyses suggest a negative affectivity factor explains covariance across PTSD, generalized anxiety, and major depression (27,28), and indicate common genetic and environmental correlates of these concerns(29).

Longitudinal studies including internalizing and PTSD symptoms have produced inconsistent results (30,31). A study of combat veterans and prisoners of war assessed for depression and PTSD over three occasions found bidirectional relationships between PTSD and depressive symptoms, suggesting a common posttraumatic construct (31). In contrast, a study of urban US civilians assessed over three occasions found PTSD and depression were related bi-directionally, but were distinct constructs (30). Other research identified gender differences in directionality, finding a bi-directional relationship among women, but unidirectional association from PTSD to depression among men (19). Another community-based study found weekly changes in PTSD over a 6 to 12-week period were predicted by pre-treatment anxiety sensitivity (32). Finally, longitudinal studies of war veterans and lung injury survivors suggest anxiety, depressive, and PTSD symptoms emerge as concurrent co-morbidities(9,33). Therefore, the current study aims to address these discrepancies by demonstrating a consistent and conserved directionality to these symptom domains.

Recent advances in machine-learning support more flexible modeling of longitudinal associations between PTSD and internalizing symptoms. Causal inference algorithms can identify the most plausible network of directional associations, supporting data-driven investigations into the relations among these symptom domains (34–36). Unlike prior studies, these algorithms can also incorporate other outcomes that could mediate relations between PTSD and internalizing symptoms, such as alcohol use or functional status. Understanding whether and how PTSD and internalizing symptoms drive health and functioning is also important.

The present study used advanced machine learning methods to investigate the directional associations among posttraumatic and internalizing symptoms along with multiple psychosocial outcomes of clinical significance. We aimed to more precisely assess the driving roles of each symptom domain through applying causal discovery techniques to longitudinal data assessing PTSD, depressive symptoms, and psychosocial outcomes over 8 years of data. We validated our findings using multiple measures of internalizing symptoms in two additional samples from longitudinal studies with distinct age, gender representation, prevalent diagnoses, and treatment settings.

## MATERIALS AND METHODS

### Overview

The current study focused on interrelations among PTSD and other internalizing symptoms using data-driven and hypothesis-driven computational experiments on complete longitudinal clinical data from three studies of PTSD symptom trajectories (Study 1, n=240; Study 2, n=79; Study 3, n = 116). These three datasets, and subsequent analyses, are conceptualized in **Figure 1**. Data-driven testing was performed on the Mind Your Heart (MYH) study(37) that includes 8 consecutive years of data measured across domains of PTSD, depression, social function, physical health and alcohol use. The data-driven results were replicated using data from studies of the Coordinated Anxiety Learning and Management (CALM)(38) intervention and of a Women’s Treatment for Trauma and Substance Use Disorders (WTTS)(39), drawn respectively from the NIMH Data Archive (NDA) and National Institute for Drug Abuse (NIDA) repositories. The PRISMA diagram presented in **Figure S1** shows the procedures used to identify each dataset, samples included at each analytic stage, and the overall analytic flow of the study.

**Figure 1.**
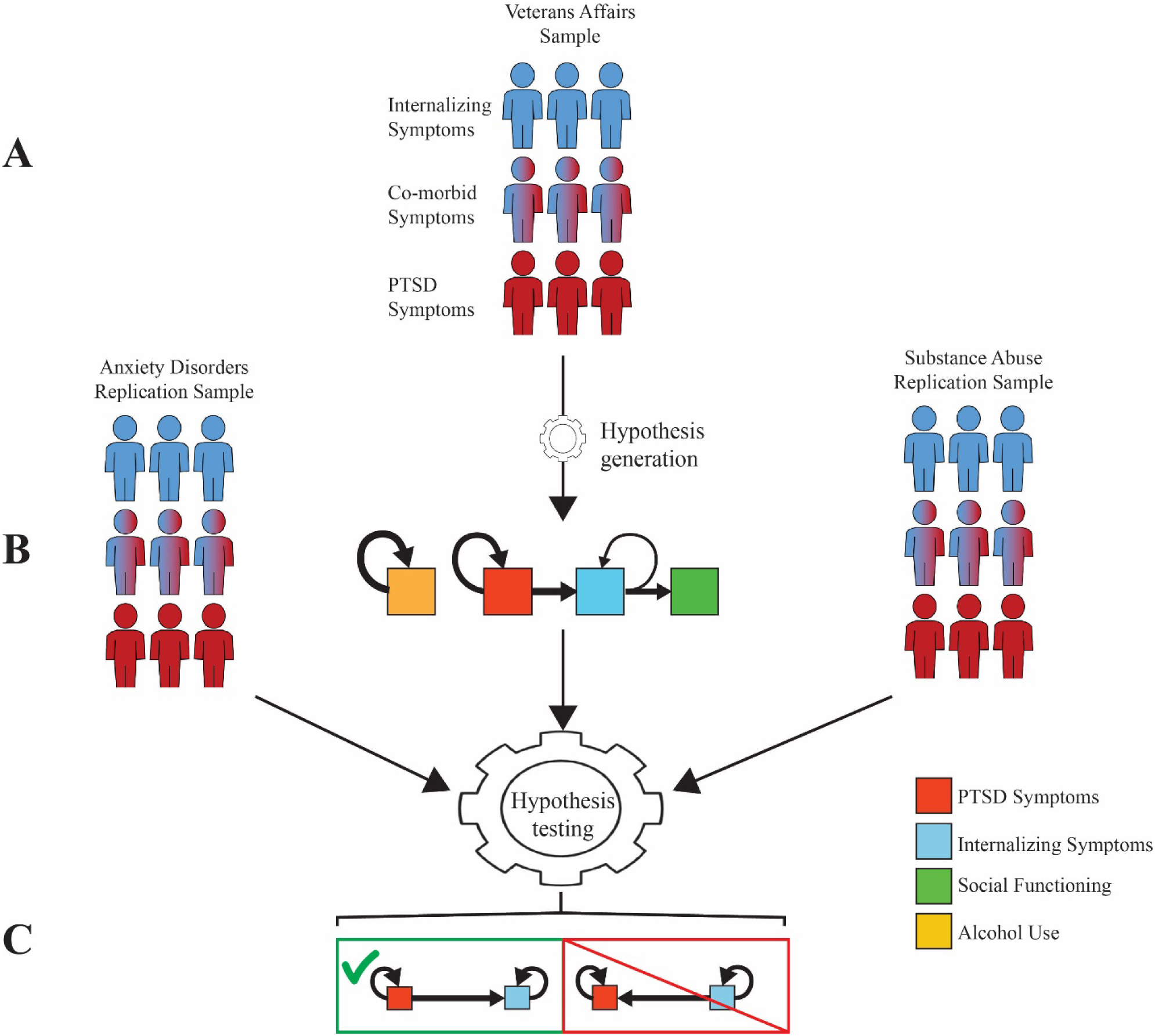
Study overview and conceptual model. A. The Mind Your Heart veterans affairs sample was used to generate an hypothesis about the relation between PTSD symptoms and internalizing symptoms in the presence of psychosocial and substance use assessments. B. A directional relation between PTSD and internalizing symptoms was identified in the hypothesis generation stage and tested on anxiety disorders and substance abuse samples. C. Competing models with PTSD symptoms driving internalizing symptoms (left) and internalizing symptoms driving PTSD symptoms (right) tested in the replication samples favored a model with PTSD symptoms driving internalizing symptoms. Recursive arrows in each graph represent autoregressive effects from prior assessments.

### Study Populations

The prospective MYH Study was designed to examine associations between PTSD and health outcomes. Participants were recruited between 2008 to 2010 from the San Francisco and Palo Alto Department of Veterans Affairs (VA) medical centers. Flyers at VA facilities and mailings were used to recruit patients with and without PTSD diagnoses in the previous 5 years(40,41).. Participants were excluded if they had a myocardial infarction 6-months prior, could not walk one block on a treadmill, did not have stable contact information, or planned to move within 3 years. The study was approved by the institutional review boards of the University of California, San Francisco and the San Francisco Veterans Administration and all participants provided written informed consent.

747 enrolled participants completed baseline examinations. Participants completed annual telephone interviews with validated assessments of several health and function domains. The present study included data from 240 participants that completed assessments across all 8 years.

The CALM study is a randomized-controlled trial of the CALM Tools for Living intervention for anxiety disorders in primary care (42). Participants were recruited from and received treatment at four U.S. primary care sites between 2006 and 2008. Eligibility criteria included age 18 years or older and meeting criteria for any DSM-IV anxiety disorder (further exclusion criteria reported in (38)). Participants were randomized using stratified permutated block randomization and received either the CALM intervention or treatment-as-usual. Participants assigned to the intervention chose between taking medications, the Tools for Living intervention, or both. The CALM Tools for Living intervention involved eight, hour-long, computerized cognitive-behavioral intervention modules that were supervised by an anxiety specialist clinician. 1004 primary care patients who completed baseline assessments were enrolled(42), and re-assessed on outcomes at 6, 12, and 18 months post-intervention. For replication analysis, we identified 79 participants meeting diagnostic criteria for PTSD and assessed on posttraumatic stress symptoms.

The WTTS study compared two manualized interventions for co-occurring substance abuse and posttraumatic stress among women.(39) Women with substance abuse disorders and PTSD, or sub-threshold posttraumatic stress, were eligible. Participants were randomly assigned to receive either Seeking Safety or Women’s Health Education interventions to supplement ongoing treatment for substance abuse. Participants were assessed on a range of outcomes at baseline, 2-months later at post-intervention, and at 3, 6, and 12-month follow-ups. Details of the WTTS study design are published elsewhere (39). Of the 353 participants initially enrolled in the WTTS study, our second replication analysis studied 116 with complete data on all measures at each assessment point.

### Measures

Each study differed in methods of measuring PTSD and internalizing symptom domains, but all used validated questionnaires. To assess PTSD symptoms, MYH and CALM used the PTSD Checklist (43) and WTTS used the PTSD Symptom Self-Report scale(44). For internalizing symptoms, MYH used the Patient Health Questionnaire(45) to assess depression; CALM used the Goldberg Anxiety and Depression Scale(45–48) to assess anxiety and depression co-morbidity as well as the Brief Symptoms Inventory - Anxiety scale to assess anxiety(49–51); and WTTS used the Brief Symptoms Inventory – Depression and Anxiety scales to assess depression and anxiety, respectively (49,51,52). Diagnostic status was assessed at baseline using the Composite International Diagnostic Interview for the DSM-IV(53) in MYH, the MINI International Neuropsychiatric Interview(54) in CALM, and the Clinician Administered PTSD Scale(55) in both WTTS and MYH for PTSD diagnoses.

Additional psychosocial outcomes were included in the data-driven analyses performed on the MYH study, to assess intervening factors in the relation between PTSD symptoms and depressive symptoms. These measures included the Alcohol Use Disorders Identification Test (AUDIT) - Consumption scale(56,57); the Short Form-36 General Health, Physical Functioning and Social functioning scales(58,59); and 5-point Quality of Life and physical activity rating scales.

### Analyses

#### Data-Driven Hypothesis Generation

Greedy Fast Causal Inference (GFCI)(34,35) analysis was performed to determine the network structure among PTSD symptoms, internalizing symptoms, and related outcomes in the MYH data. GFCI uses a combination of goodness-of-fit statistics, conditional independence tests, and artificial intelligence to search across all possible directed acyclic graphs (DAGs), including DAGs with unmeasured variables, to find the collection of DAGs most consistent with the data. DAGs are used to represent the structure of models, including Structural Equation Models (SEMs)(60) and Bayesian Networks (BNs)(61), and GFCI is analogous to searching through the space of all possible SEMs or BNs. This collection of DAGs is represented as a mixed ancestral graph (MAG). The lines connecting the nodes in MAGs can have a combination of different endpoints, e.g. arrowheads, arrow tails, and circles, along with different line types, capturing information about the entire set of represented DAGs.

The DAG space is extremely large and impossible to search exhaustively for the number of variables in the primary dataset. GFCI selects an initial set of DAGs using a depth-first search guided by artificial intelligence (AI) to optimize a comparative goodness-of-fit statistic. It then uses AI to modify these graphs to maintain consistency with conditional independence and dependence statements supported by the data. These modifications can include the addition of unobserved factors. The resulting MAG thus represents a set of DAGs which both outperform all alternative theories and are as consistent with the data as possible. This makes GFCI a robust analytic approach that maximizes information gain from highly multidimensional data without many of the shortcomings of traditional clinical prediction models.

GFCI was run using Tetrad version 6.6.0. The primary settings and parameters were left as their defaults: Bayesian Information Criterion (BIC) score with penalty discount 2, and Fisher Z test with alpha 0.01. The upper bound on maximum degree was removed by setting it to −1. This was a default value for older versions of Tetrad and prioritizes accuracy over runtime. The algorithm’s search was augmented with background knowledge to restrict causal arrows pointing backwards in time. Graph stability was assessed from 1,000 bootstrap samples, applying the same GFCI analysis to each, and aggregating the resulting 1,000 graphs into a table summarizing the proportion of all possible relationships. The edges in the graph were compared to those in the table to confirm that they were not regularly changed or destroyed.

### Effect size estimation

To obtain effect-size estimates, each MAG was converted to a path analysis input(62). Each directional relation was represented as a regression path, whereas non-directional relations in the MAGs were represented as covariances in the path model. Aside from relations among the baseline measures in each study, all covariance relations not included in the MAGs were constrained to zero in the path analysis model. The resulting models thus paralleled the robust causal network represented in the MAGs, while preserving baseline relations among variables of interest. The path models were run using the “lavaan” package in R(63) using the default maximum likelihood estimation procedure. Standardized associations and confidence intervals were interpreted to obtain effect-size and uncertainty estimates associated with relations depicted in the MAGs.

### Replication of Hypothesis

Our replication analyses tested the identified directional relationships between PTSD and other internalizing symptoms from the MYH study using comparisons of path analytic models in the CALM and WTTS datasets. These analyses included multiple measures of internalizing symptoms to determine the generalizability of the data-driven hypotheses. Models were specified using the “lavaan” package (63) and model comparisons were performed using hypothesis tests for non-nested models described in Vuong (64) via the “nonnest2” package (65) in R. Significant differences among models were described via differences in the log-likelihood and BIC fit indices estimated through “lavaan.” These indices assess how well competing models reproduced observed data, allowing for comparison between competing models on the same dataset. The best-fitting models identified through the hypothesis tests and fit indices were interpreted in relation to the hypotheses generated from the initial GFCI analyses on the MYH data.

## RESULTS

We observed several significant differences in baseline characteristics of all three cohorts. **Table 1** shows demographic and diagnostic comparisons among participants in each sample that had complete data at all time points. Age significantly differed across samples, with participants in MYH being the oldest, followed by CALM and then by WTTS. There was a larger proportion of White or European-American participants in CALM and a larger proportion of Black or African-American participants in WTTS, and no participants identifying as Asian or Pacific-Islander in WTTS. A greater proportion of participants in CALM identified as Latinx or Hispanic, as compared with the other samples. Educational background differed between MYH and CALM, with a greater proportion of CALM participants endorsing college experience (education was not available for WTTS). Rates of PTSD diagnosis differed across the full cohort samples: WTTS participants showed the highest rate (80.2%), followed by MYH participants (34.5%), and CALM participants (7.9%). Rates of major depression and generalized anxiety disorder differed across the MYH and CALM samples (**Table S1**) and were not assessed in WTTS. Additionally, comorbid PTSD and depression were more prevalent in the MYH study (39% of the cohort) versus the CALM study (15%), paralleling rates in samples of veterans (9) and treatment-seeking individuals (66). Diagnostic rates differed in the CALM analysis sample, as only participants with PTSD were included from this sample. The most notable difference related to gender: MYH participants were mostly male, the CALM study had a more even distribution between males and females, and WTTS participants were all female.

**Table 1.**
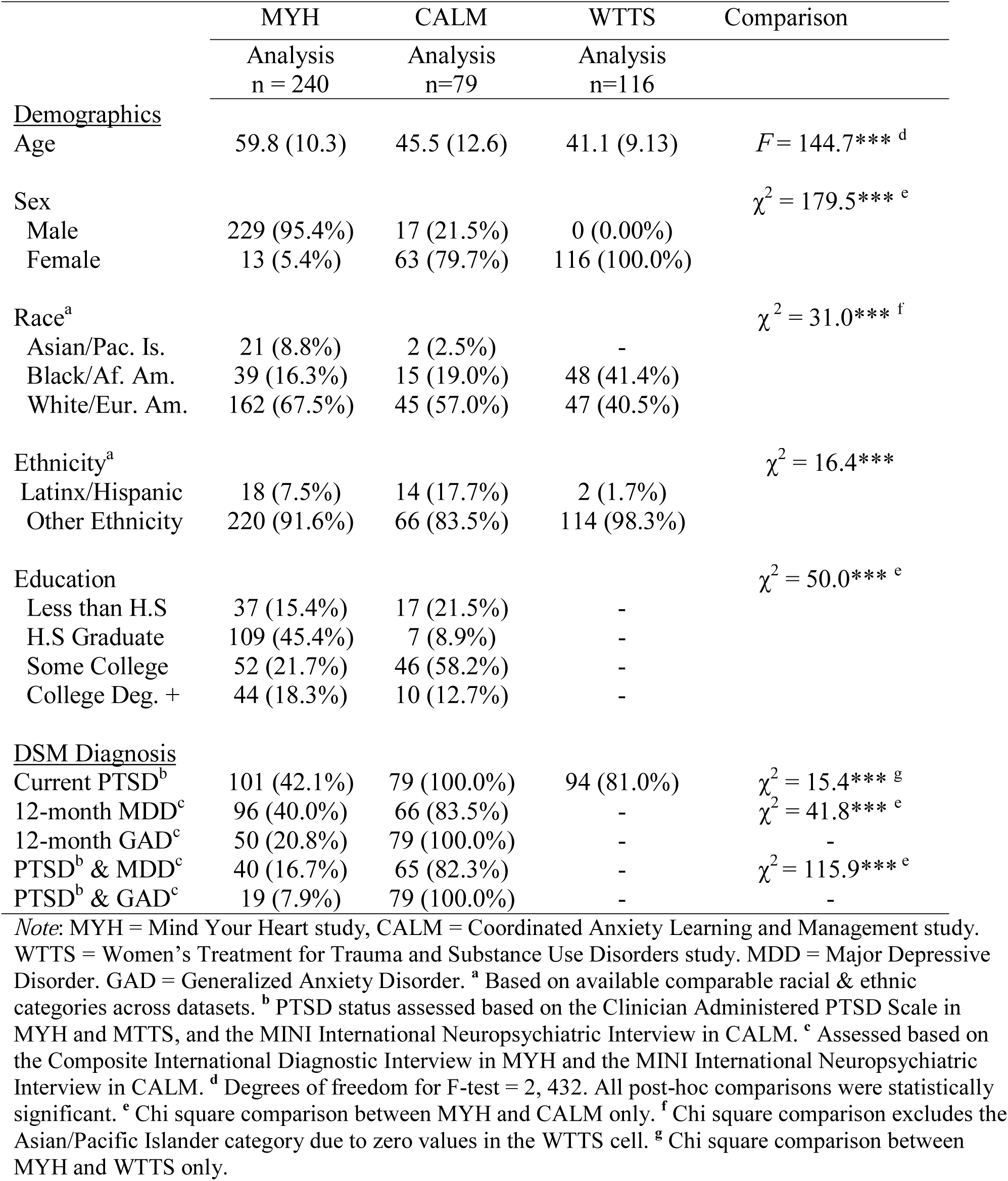
Demographics, descriptive statistics, and comparison between analysis samples from the Mind Your Heart, Coordinated Anxiety Learning and Management, and Women’s Treatment for Trauma and Substance Abuse studies.

### Data-driven hypothesis generation in the MYH study

**Figure 2** shows the pattern of relations implicating PTSD symptoms, depressive symptoms, and related measures identified most consistently across assessments. PTSD symptoms drove later PTSD symptoms, depressive symptoms, and social functioning via depressive symptoms across MYH assessments. Depressive symptoms drove later depressive symptoms on only two of the measurement occasions, while social functioning did not drive itself from one time point to the next. Meanwhile, alcohol use drove only itself and shared no edges with other measures. **Figure S2** displays the full GFCI network, including other psychosocial variables that did not interact directionally with PTSD or depressive symptoms, without rolling longitudinal arrows into a single loop. Physical function varied largely independently across assessments, driving itself but not influencing or being influenced by other domains. Similarly, quality of life and overall health appeared to drive each other.

**Figure 2.**
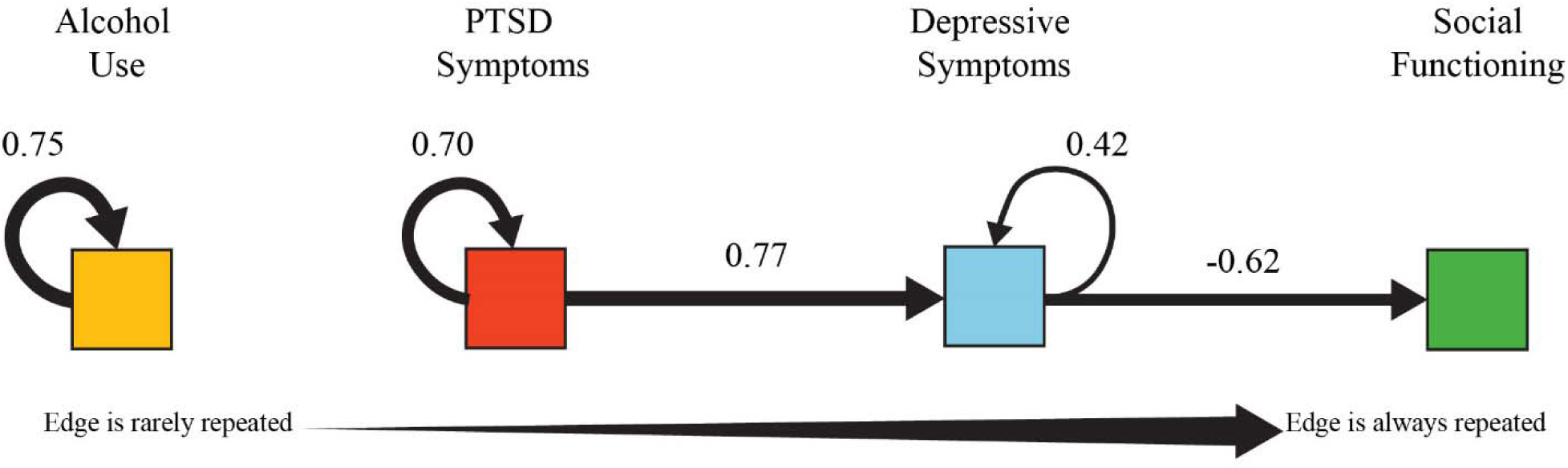
Rolled network graph of primary Greedy Fast Causal Inference (GFCI) outcomes in the Mind Your Heart dataset. Summary of the recursive pattern of relations that emerged across assessment points of the Mind Your Heart study. Arrows bridging between variables represent directional, cross-sectional paths identified through GFCI. Arrows looping recursively into the same variable represent autoregressive effects from prior assessments to later assessments on that variable (e.g. repeated measures). The thickness of each arrow represents the frequency with which the path occurred across the eight assessment points of the Mind Your Heart study.Path estimates with numerical values represent standardized effects, averaged across time-points. Per this graph, PTSD symptoms appear to be an outcome of primary importance, as it causally influences itself as well as depression and social functioning, and alcohol use is completely independent of these other symptoms. The full GFCI network and path estimates at each of the assessment points are presented in supplementary Figure S2.

The same analysis was run for only the first three MYH time points to boost the sample size to 508, and the same network was reproduced. Additionally, to test whether these results differed between participants with PTSD (n=101) or without a PTSD history (n=136), GFCI was performed again on these subgroups separately and compared (**Figure S3**), again reproducing the general structure and relationships in the network. Stability analyses of 1,000 bootstrapped samples of MYH supported the structure of relations identified in the MYH study and are reported in supplementary **Table S2**.

### Replication of network structure in CALM and WTTS datasets

Data on 79 participants from CALM and 116 participants from WTTS were used for external cross-validation of the outcome relations identified in MYH. To cross-validate the directional relation from PTSD symptoms driving internalizing symptoms, two competing and equal *df* models shown in **Figure 1C** were compared in each study. We hypothesized that models including a directed edge from PTSD symptoms to concurrent internalizing symptoms at each assessment would fit the data better than models with an edge from internalizing symptoms to PTSD symptoms. This hypothesis was borne out using both depressive and anxious symptom measures in both the CALM and WTTS samples, as displayed in **Table 2**. Vuong’s closeness tests indicated significantly better fit of the PTSD-driven models compared with internalizing-driven models across all samples. Differences in BIC indicated a large decline in fit from the PTSD-driven models to the internalizing-driven models. Supplementary **Figures S4** and **S5** display path models for the competing hypotheses tested in CALM and WTTS and their respective BIC. The standardized paths from PTSD symptoms to internalizing symptoms were consistent in magnitude with those observed in the MYH sample.

**Table 2.** Model fit information & comparisons for replication analyses.

| <u>Sample Measures</u><br>Fit Index | PTSD Sx →<br>Internalizing Sx | Internalizing Sx<br>→ PTSD Sx | Difference |
| --- | --- | --- | --- |
| <u>CALM</u> |  |  |  |
| *PCL & GADS* |  |  |  |
| Log Likelihood | -1732.5 | -1820.0 | Vuong Z = 11.95*** |
| Bayesian Information Criterion | 3539.2 | 3714.3 | ΔBIC = 175.1 |
| *PCL & BSI-A* |  |  |  |
| Log Likelihood | -2001.5 | -2037.4 | Vuong Z = 4.15*** |
| Bayesian Information Criterion | 4077.3 | 4149.1 | ΔBIC = 71.8 |
| <u>WTTS</u> |  |  |  |
| *PSSR & BSI-D* |  |  |  |
| Log Likelihood | -2200.2 | -2486.0 | Vuong Z = 28.08*** |
| Bayesian Information Criterion | 4505.0 | 5076.6 | ΔBIC = 571.6 |
| *PSSR & BSI-A* |  |  |  |
| Log Likelihood | -2130.9 | -2415.4 | Vuong Z = 32.80*** |
| Bayesian Information Criterion | 4366.4 | 4935.3 | ΔBIC = 568.9 |

*Note.* PCL = PTSD Checklist. GADS = Goldberg Anxiety and Depression Scale. BSI-A = Brief Symptoms Inventory – Anxiety. PSSR = PTSD Symptoms Self-Report. BSI-D = Brief Symptoms Inventory – Depression. $\Delta$ BIC = Difference in BIC fit index between models, with $\Delta$ BIC > 10 reflecting very strong evidence against the worse-fitting model. Vuong Z = Likelihood-based comparison among models, distributed as Z-statistic. \*\*\*Vuong Z is significant at $p < .001$ .

### Exploratory analysis of potential mechanisms

An additional exploratory GFCI analysis was performed to examine the relations among PTSD symptom clusters and depressive symptoms in the MYH sample to identify possible mechanisms of this relationship. As items of the PTSD Checklist (PCL) in MYH were formulated for the DSM-IV, we re-categorized them according to DSM-V re-experiencing, avoidance, negative alterations in cognition and mood (NACM), and hyperarousal clusters (see (67) for a previous analysis taking this approach). Multilevel confirmatory factor analysis (MLCFA) was fitted to the items, with four factors representing the DSM-V symptom clusters (see **Figure S6** and the Supplementary MLCFA Method and Results). Finally, scores for each symptom cluster were computed, and GFCI analyses were used to explore relations between these symptom clusters and depressive symptoms over time. The first GFCI analysis revealed a strong relation between NACM and depressive symptoms (**Figure 3A**), likely due to strong similarities in symptom domains included in NACM and depression as assessed by the PCL and Patient Health Questionnaire. Therefore, we removed the NACM cluster in a second GFCI, which revealed hyperarousal as the primary driver of depressive symptoms as well as the other PTSD symptom clusters (**Figure 3B**). Taken together, these results suggest NACM is most proximally related to depressive symptoms, while hyperarousal may be a central factor driving these symptoms across time. **Figure S7** provides a detailed depiction of the full GFCI network including the DSM-V PTSD symptom clusters and depressive symptoms in the MYH data.

**Figure 3.**
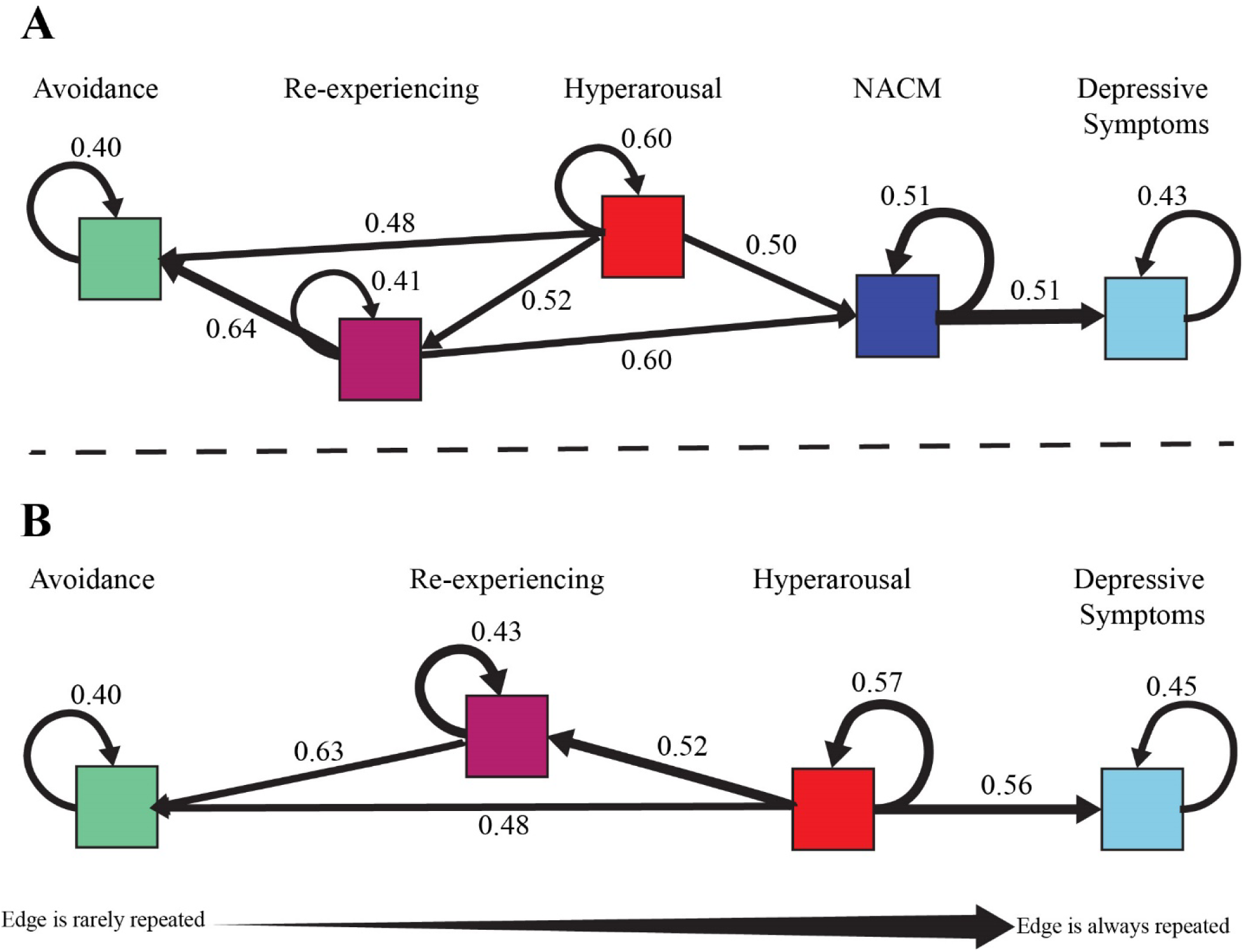
Rolled network graph of exploratory Greedy Fast Causal Inference (GFCI) analysis of potential mechanisms. Summary graph of the recursive pattern of relations that emerged in the GFCI analyzing DSM-V symptom clusters and depression scores in the Mind Your Heart study. A. Rolled graph showing recurring paths across measurement occasions for the GFCI analysis on the DSM-V PTSD symptom clusters and depressive symptoms including NACM. B. Rolled graph for the GFCI analysis on the DSM-V PTSD symptom clusters and depressive symptoms excluding NACM. NACM bridged between other PTSD symptoms and depressive symptoms when it was included in the model (A), whereas hyperarousal bridged between the other PTSD symptoms and depressive symptoms when NACM was removed (B). All paths presented occurred on least two assessments. The full GFCI network and path estimates at each of the assessment points are presented in supplementary Figure S7. DSM-V = Diagnostic and Statistical Manual V. NACM = Negative Alterations in Cognition and Mood.

## DISCUSSION

This is the first study to use a data-driven approach to identify and replicate a unidirectional relationship from PTSD to internalizing symptoms across multiple, distinct samples with varying diagnostic and demographic characteristics. Our findings provide insight into trajectories of patients with co-morbid diagnoses, with implications for treating internalizing symptoms in the context of PTSD. These results highlight the importance of measuring trauma exposure and presence of PTSD in clinical research studies of internalizing disorders and controlled clinical trials for internalizing distress.

Studies evaluating associations of PTSD and internalizing symptoms over time have had conflicting results (9,17,27-30). This research has found unidirectional relations between PTSD and internalizing symptoms (though directionality was not consistent, 17, 29); bi-directional associations(9,27,28,30); and differential associations by population characteristics (e.g., gender)(17). We expand on this work by using machine learning methods that offer more flexible analyses of the associations among PTSD and internalizing symptoms, along with potential confounders. Previous research relies on testing competing *a-priori* hypotheses for these relations; for instance, by specifying competing models and then assessing their correspondence to the data (27,28). This research is often limited to a single sample with few assessment points across which to establish conserved symptom relations (19-23, 27-29). In contrast, the present study used a data-driven approach that allowed for nuanced relations to emerge across multiple timepoints in a hypothesis generation stage, and then reproduced across two additional samples. Our finding of a unidirectional association with PTSD explaining concurrent internalizing symptoms, after accounting for prior internalizing symptoms, was consistent across three cohorts representing separate studies of primarily men or women, and observational and clinical trial samples across both veteran and civilian cohorts. Beyond these results, our mechanistic analyses suggested a bridging role of hyperarousal and cognitive and affective disruption between PTSD and depressive concerns more specifically.

Our findings raise questions about the interpretation of internalizing symptoms that emerge in the context of PTSD. Such symptoms may not be accurately characterized as involving co-morbidity among distinct conditions. Instead, our results suggest these symptoms represent a downstream outcome of PTSD, which may be addressed through treating features bridging between posttraumatic stress and internalizing concerns. Consistent with our exploratory analysis of potential mechanisms, a study of Israeli veterans suggested centrality of the DSM-V NACM symptom cluster in bridging among PTSD, depressive symptoms, and moral injury (25). Similarly, consistent with our findings on the centrality of hyperarousal across PTSD and depressive symptom groups, other research suggests anxiety sensitivity and avoidance of inner experiences broadly may be transdiagnostic processes underlying PTSD and internalizing symptomatology (32,68,69). Further longitudinal research should consider the mechanisms connecting hyperarousal, NACM, and other internalizing symptoms that could play a transdiagnostic role in comorbidities.

Our findings affirm the need to assess for posttraumatic stress in the context of research on anxious and depressive disorders. This appears especially pressing for clinical trials, as there is a lack of information about the co-occurrence of PTSD in much of this research (18,66). For example, we were unable to find a study with a focus on internalizing symptoms and sufficient sample size that also assessed for PTSD across the NDA and NIDA repositories. This issue may be less a result of actively excluding patients with co-morbidities, but more a product of limited assessments of co-morbid symptomology in such studies (70). Conversely, studies may exclude more severe or sub-threshold manifestations of either PTSD or internalizing symptoms (71), inadvertently restricting information about possible co-morbid conditions. A lack of information about co-morbidity may drive inaccurate care when a trauma-informed approach is not taken, and may neglect crucial mechanisms that could support clients’ recovery.

While our primary analyses concerned the relation between PTSD and internalizing symptoms, our exploratory analyses generated other hypotheses pertaining to PTSD and psychosocial outcomes. Specifically, our GFCI analyses on the MYH cohort suggested social functioning is indirectly affected by PTSD symptoms through depressive symptoms. This finding aligns with studies showing that treating interpersonal problems in PTSD can ameliorate comorbid major depression(72,73). Conversely, our findings suggest changes in PTSD symptoms provide little information about changes in alcohol use, physical functioning, and overall health over time, and vice versa. Individuals with alcohol use or physical health concerns may thus benefit from combined treatment approaches. The alcohol use findings are noteworthy as previous studies have documented co-morbidities among PTSD and alcohol abuse, yet rarely examine whether PTSD symptoms drive alcohol abuse longitudinally (74). These findings are important to replicate in future research integrating PTSD symptoms and psychosocial outcomes.

Our results are subject to several limitations. While GFCI can infer ‘causality’ from repeated measures based on statistical criteria, the MYH study was observational. Typically, causality is confirmed with data from randomized controlled trials. We cannot randomize patients to develop psychiatric disorders, but machine learning techniques can be applied to repeated measures of PTSD, internalizing symptoms, and psychosocial outcomes collected during intervention trials to overcome this. Our validation cohorts were drawn from clinical trials, but only a subset of participants had both PTSD and internalizing symptoms assessed. Therefore, replication with thorough measurement of both symptom domains will be important. Second, though we conducted our analyses in samples of largely male veterans, women with substance use disorders, and a subset of patients enrolled in an anxiety trial, validation in populations with diversity in other experiences and identities is crucial. Third, patients were excluded due to missing data on repeated assessments, although findings based on fewer time-points and excluding fewer patients were consistent with those on all time-points. Finally, our samples largely represented patients with PTSD, so replication with other salient clinical concerns would be valuable.

Despite these limitations, our finding that PTSD symptoms drove internalizing symptoms in three separate studies suggests an etiological connection from trauma symptomology to internalizing distress. This supports prior work suggesting evolution of other internalizing symptoms secondary to PTSD could represent a manifestation of the original response to traumatic stress (7,9,28). Our results underscore the need for integrated therapies that treat mechanisms underlying a constellation of patients’ symptoms, rather than addressing single disorders. Finally, this work highlights the importance of longitudinal data incorporating simultaneous assessment of PTSD, internalizing distress, and patient-centered outcomes in both observational and trial settings.

## Supporting information

Supplemental Methods, Results, and Code

## Data Availability

Data from the Mind Your Heart study is not publicly available, however details on the study design and data availability can be found at https://mindyourheartstudy.ucsf.edu/.

## Author contributions

JLN, EK, SM, ARF, TCN, BEC conceived of the study. BEC and TCN collected or oversaw collection of MYH primary data. JLN, TK and BP performed data query and wrangling of MYH and NDA datasets. EK performed GFCI on datasets. JLN, EK, BP, SM, ARF, TCN and BEC were involved in interpreting results. JLN, EK, SM, TK, BP, ARF, TCN, BEC wrote the manuscript. The funders had no role in the design and conduct of the study; data collection, management, analysis, interpretation, or manuscript preparation.

## Data and materials availability

The information reported here results from secondary analyses of data from clinical trials conducted by NIMH and NIDA, housed in public repositories, including the NIMH Data Archive (NDA: https://nda.nih.gov/) and the NIDA Data Share website (https://datashare.nida.nih.gov/). Specifically, data from the Coordinated Anxiety Learning and Management (CALM) (Study #2146 in NDA) and Women’s Treatment for Trauma and Substance Use Disorders (NIDA-CTN-0015) were included. Additional data from the Mind Your Heart study is available upon request from the Principal Investigator, Beth Cohen (https://mindyourheartstudy.ucsf.edu/contact-us). Code used in this study is available in the Code Supplement.

