## Supplemental Methods, Results, and Code for "Causal discovery identifies posttraumatic stress as a driver of internalizing symptoms across independent veteran and civilian populations"

**Table of Contents**

Supplementary Tables & Figures...…………………………………………………………	1

Figure S1. PRISMA Diagram……………………………………………………...	1

Table S1. Full Sample Descriptives………………………………………………..	2

Figure S2. Mind Your Heart GFCI analysis……………………………………….	3

Figure S3. GFCI Analysis by PTSD Diagnosis……………………………………	5

Figure S4. External validation on CALM sample………………………………….7

Figure S5. External validation on WTTS sample………………………………….	9

Supplementary Multilevel Confirmatory Factor Analysis Method…………………………11

Supplementary Multilevel Confirmatory Factor Analysis Results………………………....	12

Figure S6. Multilevel CFA for DSM-V Rescaling of the PTSD Checklist……......	13

Figure S7. GFCI Analysis on DSM-V Symptom Cluster Scores………………….	14

Supplementary References…………………………………………………………………	15

Supplementary Code………………………………………………………………………..	16

R Code for Converting Tetrad DAG Output to Path Analysis………….................	17

R Code for DSM-V Multilevel CFA on the PTSD Checklist…...............................	22

*Note.* Table S2 is appended as a separate file due to length.

**Supplementary Tables and Figures**

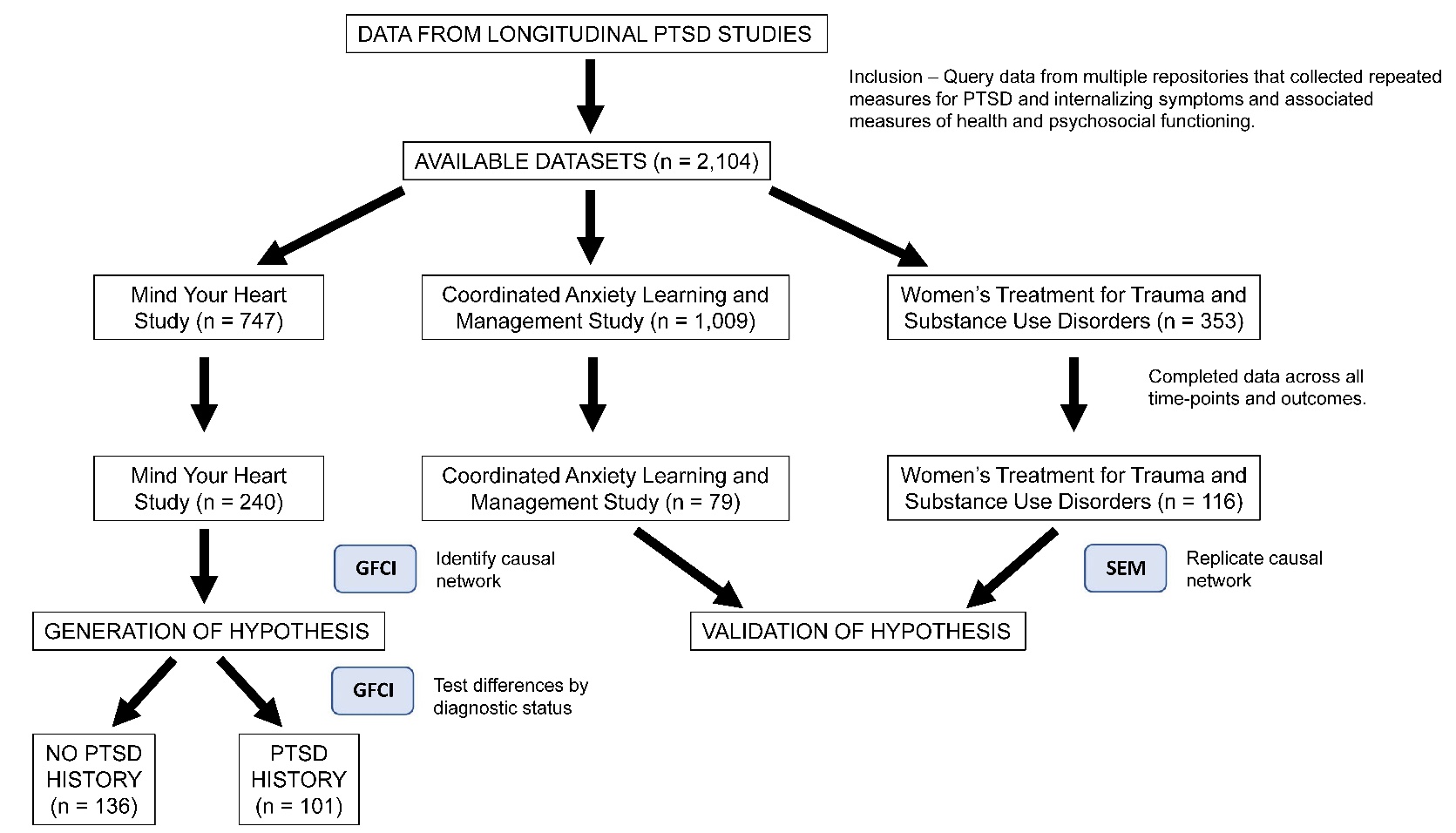

**Figure S1. PRISMA diagram showing search and selection of data for initial testing and external validation.** Data from completed longitudinal clinical studies assessing symptoms of PTSD, internalizing symptoms, and related psychosocial and health measures were identified by querying data from the Mind Your Heart (MYH) study, the NIMH Data Archive (NDA), and the National Institute for Drug Abuse data sharing repository (NIDA). One study from NDA met inclusion criteria, with approximately 10% of those subjects assessed on both PTSD and internalizing symptoms. One study from NIDA met inclusion criteria, with approximately 33% having complete observations on PTSD and internalizing symptoms.  Data from MYH were analyzed first using Greedy Fast Causal Inference methods (GFCI) to generate the initial hypothesis, which was internally tested for differences between PTSD vs no PTSD groups in MYH. Data from the CALM and WTTS studies were then used for independent validation of the causal structure identified in the MYH sample via Structural Equation Modeling (SEM) to test differences between PTSD-driven and internalizing-driven structures.

**Table S1.** Demographics, descriptive statistics, and comparisons between the full Mind Your Heart, Coordinated Anxiety Learning and Management, and Women’s Treatment for Trauma and Substance Use Disorders study samples.

|  | MYH | CALM | | WTTS | | Comparison |
| --- | --- | --- | --- | --- | --- | --- |
|  | All  n = 747 | All  n=1004 | | All  n=353 | |  |
| Demographics |  |  | |  | |  |
| Age | 58.4 (11.3) | 43.5 (13.4) | | 39.16 (9.28) | | *F* = 442.3*** ^d^ |
| Sex |  |  | |  | | χ^2^ = 742.3*** ^e^ |
| Male | 703 (94.1%) | 290 (28.9%) | | 0 (0.00%) | |  |
| Female | 44 (5.9%) | 714 (71.1%) | | 353 (100.0%) | |  |
| Race^a^ |  |  | |  | | χ^2^ = 144.5*** ^f^ |
| Asian/Pac. Is. | 65 (8.7%) | 22 (2.2%) | | - | |  |
| Black/Af. Am. | 160 (21.4%) | 69 (6.9%) | | 121 (34.3%) | |  |
| White/Eur. Am. | 432 (57.8%) | 672 (66.9%) | | 171 (48.4%) | |  |
| Ethnicity^a^ |  |  | |  | | χ^2^ = 59.4*** |
| Latinx/Hispanic | 56 (7.5%) | 185 (18.4%) | | 23 (6.5%) | |  |
| Other Ethnicity | 679 (90.9%) | 818 (81.6%) | | 330 (93.5%) | |  |
| Education |  |  | |  | | χ^2^ = 174.9*** ^e^ |
| Less than H.S | 27 (3.6%) | 223 (22.2%) | | - | |  |
| H.S Graduate | 129 (17.3%) | 67 (6.7%) | | - | |  |
| Some College | 371 (49.7%) | 351 (35.0%) | | - | |  |
| College Deg. + | 218 (29.2%) | 360 (35.9%) | | - | |  |
| DSM Diagnosis |  |  | |  | |  |
| Current PTSD^b^ | 258 (34.5%) | 79 (7.9%) | | 283 (80.2%) | | χ^2^ = 673.1*** |
| 12-month MDD^c^ | 189 (25.3%) | 647 (64.4%) | | - | | χ^2^ = 247.6*** ^e^ |
| 12-month GAD^c^ | 104 (13.9%) | 755 (75.2%) | | - | | χ^2^ = 622.7*** ^e^ |
| PTSD^b^ & MDD^c^ | 108 (14.5%) | 154 (15.3%) | | - | | χ^2^ = 0.3 ^e^ |
| PTSD^b^ & GAD^c^ | 71 (9.5%) | 79 (7.9%) | | - | | χ^2^ = 1.5 ^e^ |

*Note.* MYH = Mind Your Heart study. CALM = Coordinated Anxiety Learning and Management study. WTTS = Women’s Treatment for Trauma and Substance Use Disorders study. MDD = Major Depressive Disorder. GAD = Generalized Anxiety Disorder. **a** Based on available comparable racial & ethnic categories across datasets. **b** PTSD status assessed based on the Clinician Administered PTSD Scale in MYH and WTTS, and the MINI International Neuropsychiatric Interview in CALM. **c** Assessed based on the Composite International Diagnostic Interview in MYH and the MINI International Neuropsychiatric Interview in CALM. **d** Degrees of freedom for F-test = 2, 2101. All post-hoc comparisons were statistically significant. **e** Chi square comparison between MYH and CALM only. **f** Chi square comparison excludes the Asian/Pacific Islander category due to zero values in the WTTS cell.

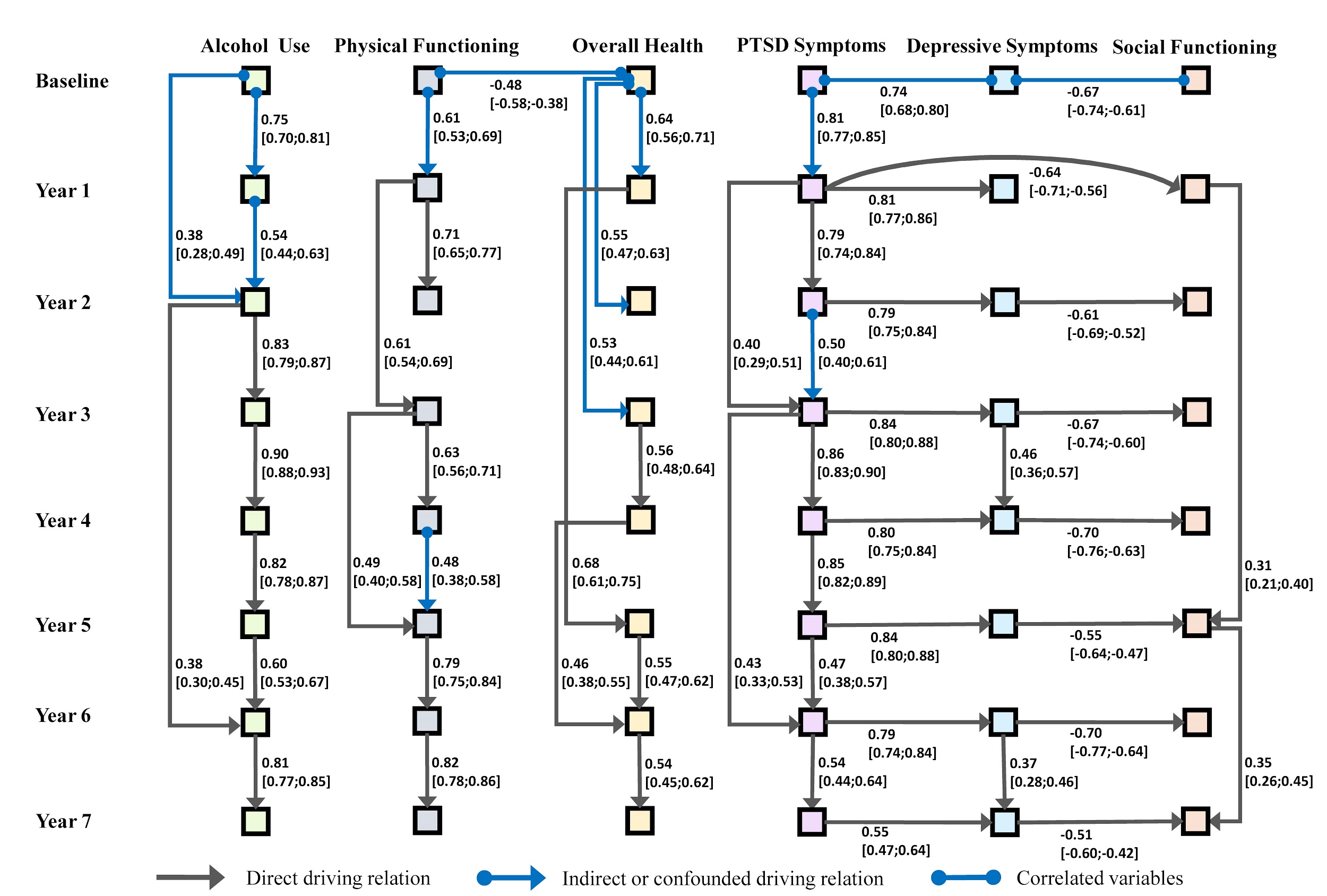

**Figure S2. Network graph analysis using Greedy Fast Causal Inference (GFCI).** GFCI was used to determine the causal structure among the outcome variables collected from baseline up until year 7 of the Mind Your Heart (MYH) study. When including all measured time points, and omitting subjects that had any missing values, 240 subjects remained. Most default parameters were selected, but maximum degree of nodes was left unbounded. Background knowledge regarding time order was used to restrict the output to graphs that do not include any causal arrows pointing backwards in time. A preliminary analysis of this causal graph at the structural level reveals: 1) PTSD symptoms appear to be an outcome of primary importance, as it causally influences itself as well as depression and social functioning. 2) Certain outcomes do not drive their own values from one time point to the next, specifically depression and social functioning. 3) Problematic alcohol use is independent of all other outcomes. 4) The same analysis was run for only the baseline, year 1, and year 2 time points, with an increased sample size of 508. The structural features previously enumerated were still present. Path analysis estimates reflect standardized regression effects, with all paths statistically significant at p < .001.

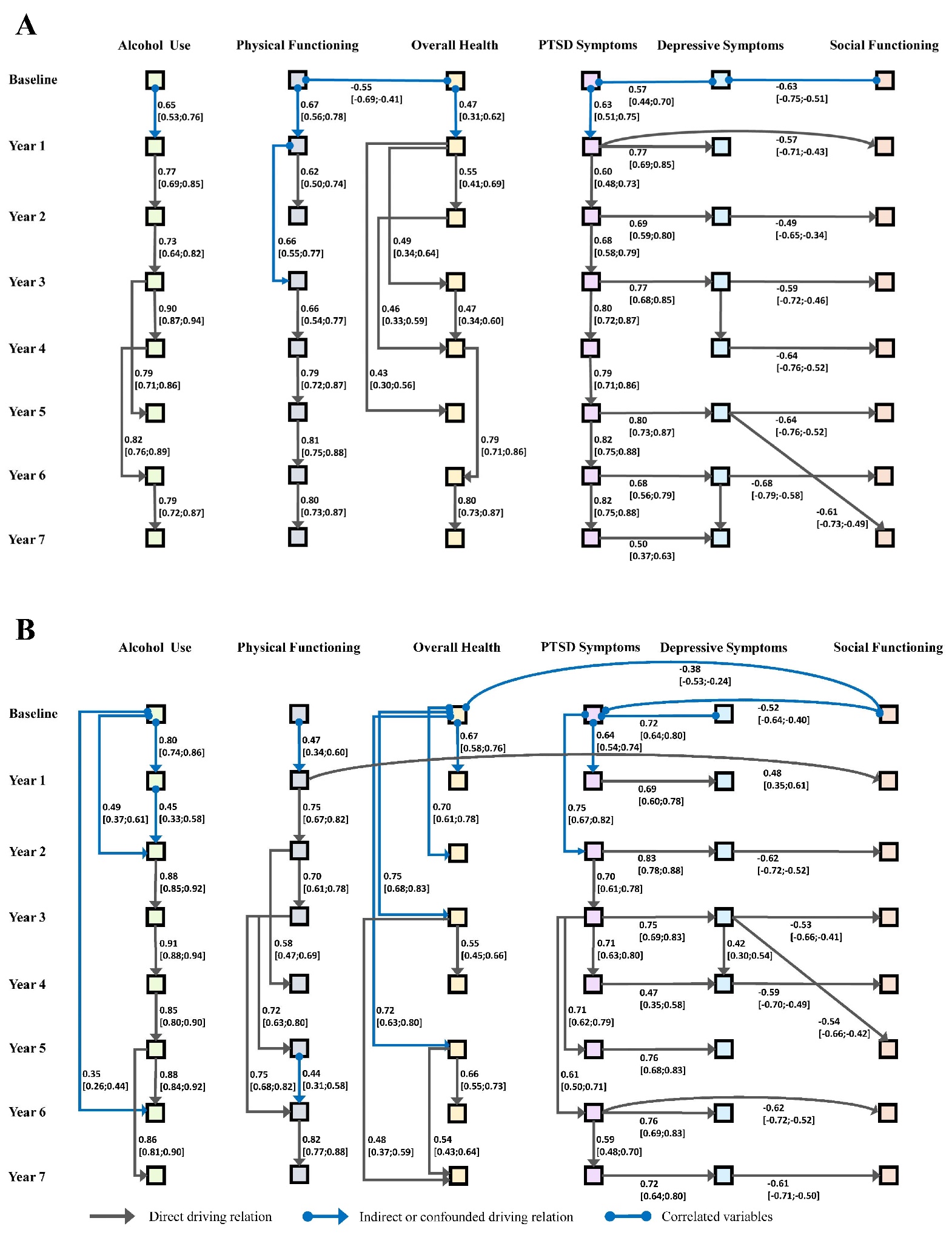

**Figure S3. GFCI performed on data from MYH study, split based on a history of PTSD or no history of PTSD.** **A** = PTSD history. **B** = No PTSD history. Structures show similarity with the entire cohort from Figure S2. A potential difference between PTSD and No PTSD is the driver relationship of physical limitation on social function at year 1 in the No PTSD group, whereas this is not the case in those with PTSD. Path analysis estimates reflect standardized regression effects. Paths statistically significant at p < .001 if bold.

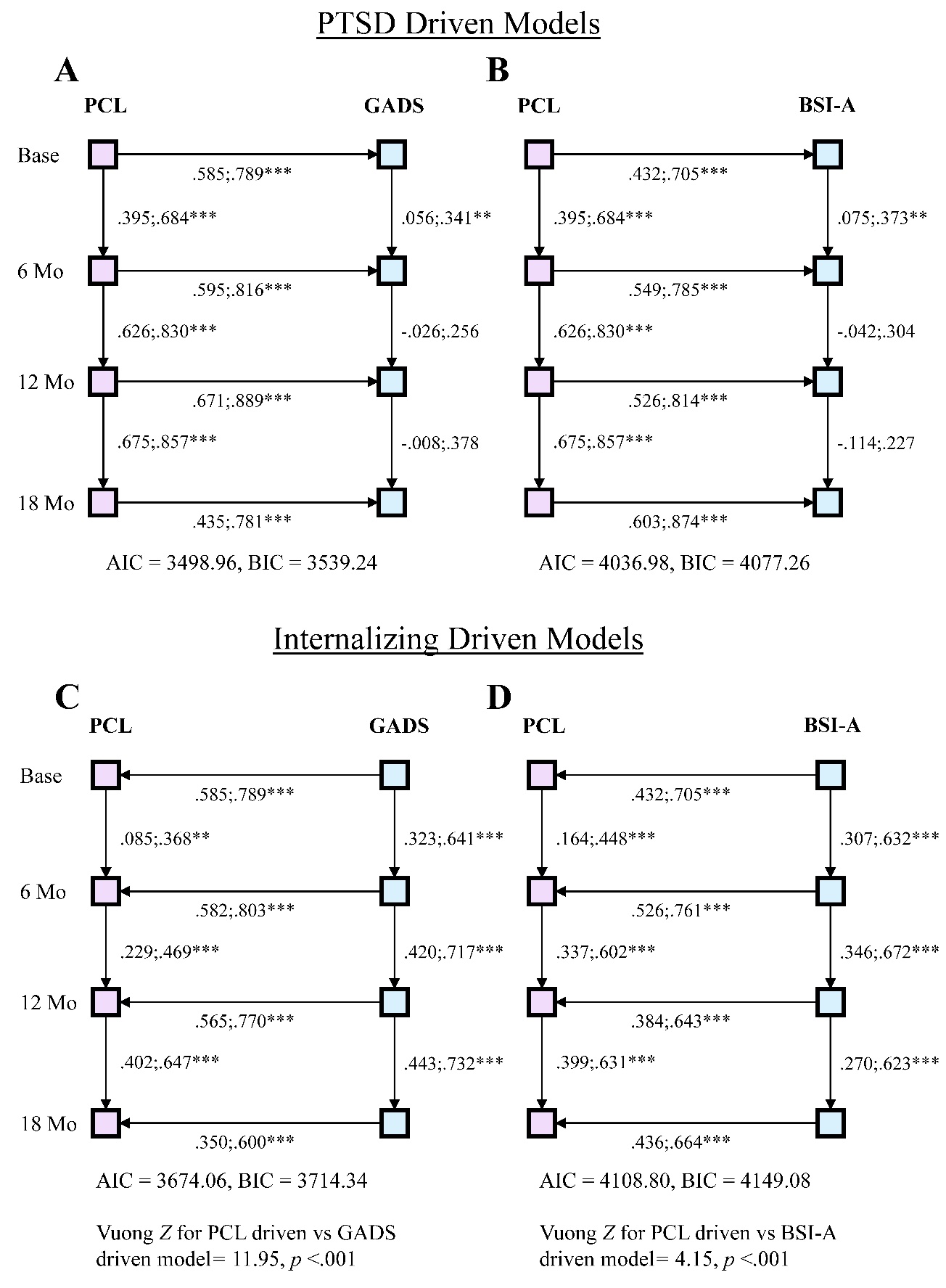

**Figure S4. External validation using structured path analytic hypothesis tests on the CALM study.** Replication of hypothesis that concurrent PTSD symptoms drive internalizing symptoms using the PTSD Checklist (PCL) to assess PTSD and the Goldberg Anxiety and Depression Scale (GADS) and Brief Symptoms Inventory – Anxiety (BSI-A) to assess internalizing symptoms. Findings indicated models with directed paths from PTSD symptoms to internalizing symptoms (A and B) fit better than models with directed paths from the internalizing measure to PTSD symptoms (C and D). Paths presented as 95% confidence intervals around the standardized effect. **p* <.05, ***p* <.01, and ****p*<.001. Vuong *Z* = Vuong’s closeness test for non-nested models. AIC = Akaike Information Criterion. BIC = Bayesian Information Criteria fit statistics. Larger AIC and BIC values indicate poorer model fit.

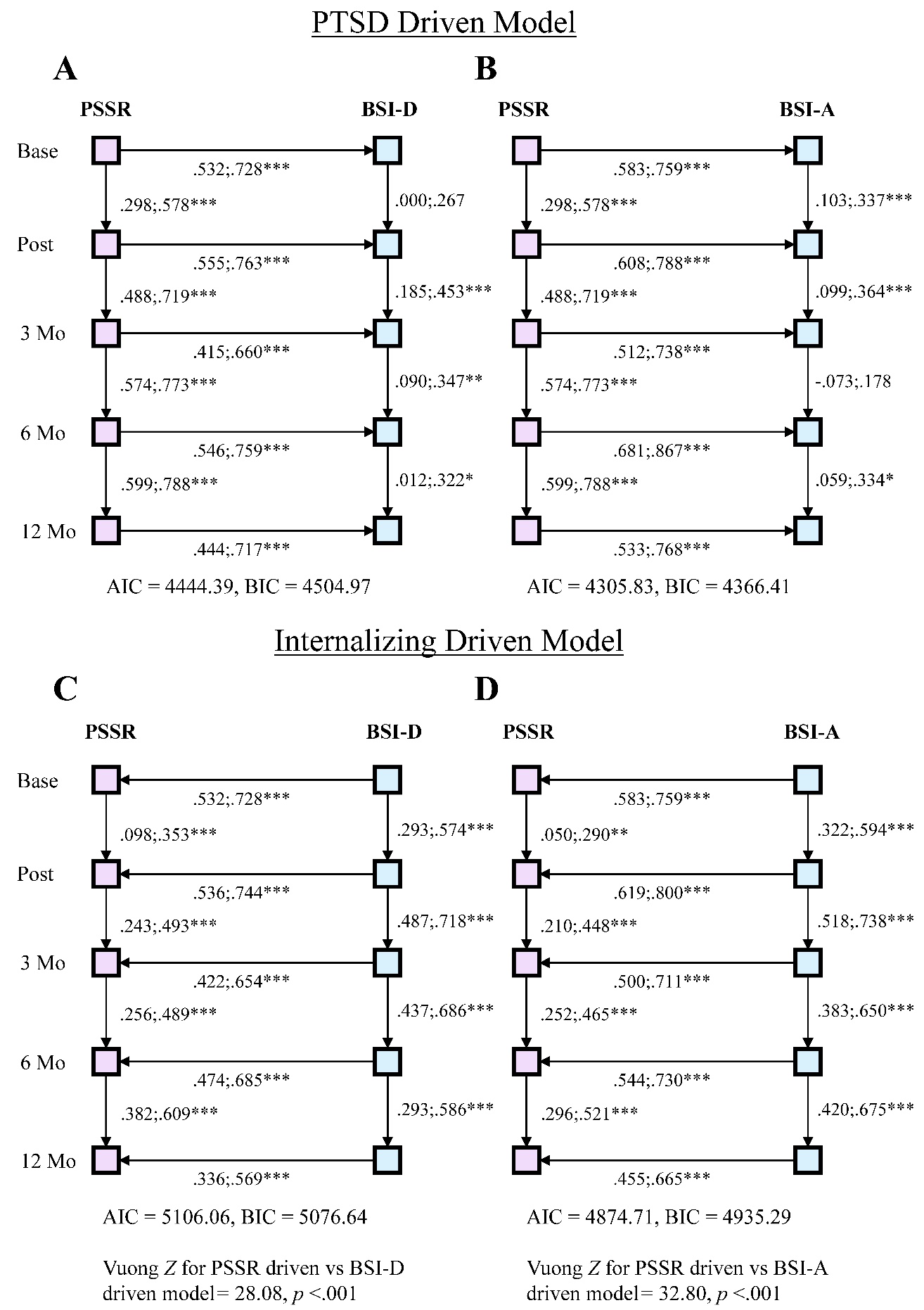

**Figure S5. External validation using structured path analytic hypothesis tests on the WTTS study.** Replication of hypothesis that concurrent PTSD symptoms drive internalizing symptoms using the PTSD Checklist (PCL) to assess PTSD and the Brief Symptoms Inventory - Depression (BSI-D) and Brief Symptoms Inventory – Anxiety (BSI-A) to assess internalizing symptoms. Findings again indicated models with directed paths from PTSD symptoms to internalizing symptoms (A and B) fit better than models with directed paths from the internalizing measure to PTSD symptoms (C and D). Paths presented as 95% confidence intervals around the standardized effect. **p* <.05, ***p* <.01, and ****p*<.001. Vuong *Z* = Vuong’s closeness test for non-nested models. AIC = Akaike Information Criterion. BIC = Bayesian Information Criteria fit statistics. Larger AIC and BIC values indicate poorer model fit.

**Supplementary Multilevel Confirmatory Factor Analysis Method**

Multilevel Confirmatory Factor Analysis (MLCFA) was used to fit a measurement model corresponding to the DSM-V PTSD symptom categorization to the PTSD Checklist items. By combining multilevel and confirmatory factor analytic frameworks, this analysis allowed for the identification of a measurement structure that would fit the data both within-persons over time and between persons (1,2). Using a multilevel parameterization, the variance of each item of the PTSD checklist was parsed into within-person variability and between-persons variability, accounting for dependencies among model residuals introduced through the nesting of repeated assessments within each person. Parallel confirmatory factor analytic measurement models were fitted to each level, and adjusted based on global fit indices such as the χ^2^ goodness of fit statistic, root mean squared error of approximation (RMSEA), comparative fit index (CFI), and Standardized Root Mean Square Residual (SRMR) as well as localized metrics of fit such as individual item residuals and modification indices describing the contribution of each parameter to the overall χ^2^ statistic (3,4). The purpose of the MLCFA was to identify a good-fitting measurement model with consistency across the within-person and between-person levels, such that each DSM-V symptom cluster would be represented by an adequate number of items with relatively equal contributions to that cluster. All MLCFA models were run using the “lavaan” package in R, using maximum likelihood estimation with robust standard errors (“MLR”) to account for departures from normality due to the ordinal (i.e., 5-point) scaling of the items (4,5).

**Supplementary MLCFA Results**

The initial MLCFA model including all 17 PTSD checklist items fit the data marginally, with χ^2^ (226) = 12304.96, *p <.*001, CFI = 0.87, RMSEA = 0.06, SRMR within = 0.07 and SRMR between = 0.04. Items 8 (trouble remembering important parts of the trauma), 9 (loss of interest in activities), 13 (trouble sleeping), and 15 (difficulty concentrating) were then removed due to modification indices suggesting strong cross-loadings onto the other factors, and due to the conceptual overlap between three such symptoms and those assessed through the internalizing symptom scale in Mind Your Heart (i.e., the Patient Health Questionnaire). The remaining items demonstrated a good fit to a four-factor structure after freeing the loading of item 3 to vary across-levels of the model and permitting correlation between the residuals of items 16 and 17, with χ^2^ (132) = 3703.83, *p <.*001, CFI = 0.96, RMSEA = 0.04, SRMR within = 0.04 and SRMR between = 0.02. This same model was then tested on the PTSD Symptoms Self-Report items in the Women’s Treatment for Trauma and Substance Abuse Disorders sample and showed adequate fit following the addition of a correlation between items 1 and 4, with χ^2^ (130) = 346.80, CFI = 0.95, RMSEA = 0.05, SRMR within = 0.05 and SRMR between = 0.07.

The results of the final MLCFA in the Mind Your Heart dataset are shown in Figure S6. Based on this model, the items loading onto each of the factors were summed to compute DSM-V PTSD symptom cluster scores. These symptom cluster scores were then entered into the exploratory Greedy Fast Causal Inference analyses along with participants’ Patient Health Questionnaire total scores to examine the longitudinal relations between participants DSM-V PTSD symptom cluster scores and depressive symptom scores. The results of these analyses are presented in Figure S7.

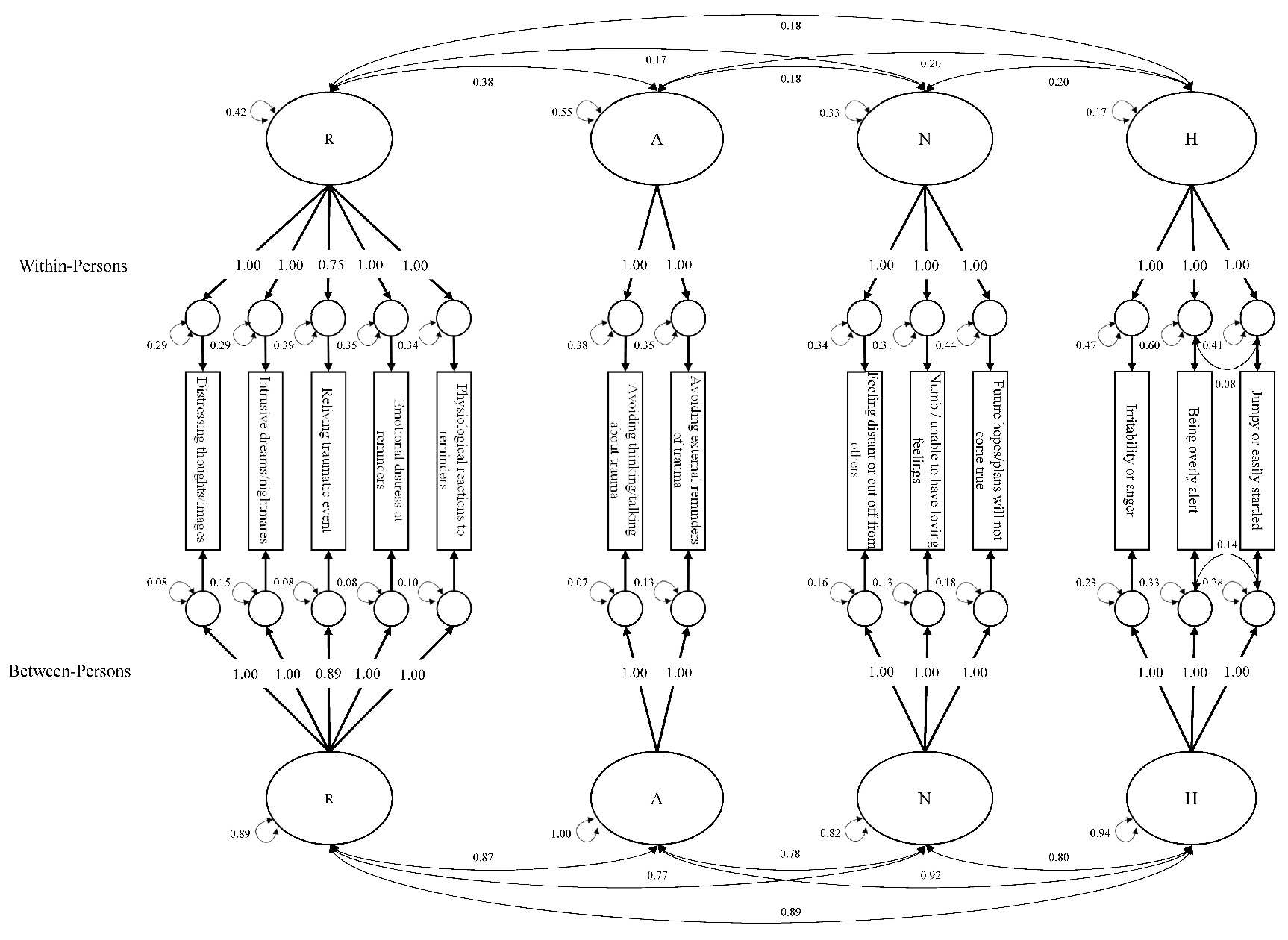

**Figure S6. Multilevel confirmatory factor analysis to assess DSM-V re-scaling of the PTSD Checklist items.** Multilevel confirmatory factor analysis (MLCFA) was used to fit a four-factor model corresponding to the DSM-V symptom clusters B through E to the PTSD Checklist items in the Mind Your Heart sample. Parallel factors of re-experiencing (R), avoidance (A), negative alterations in cognition and mood (N), and hyperarousal (H) were fit to the within- and between-person components of variance. All loadings, variances, and co-variances are statistically significant at *p* < . 001.

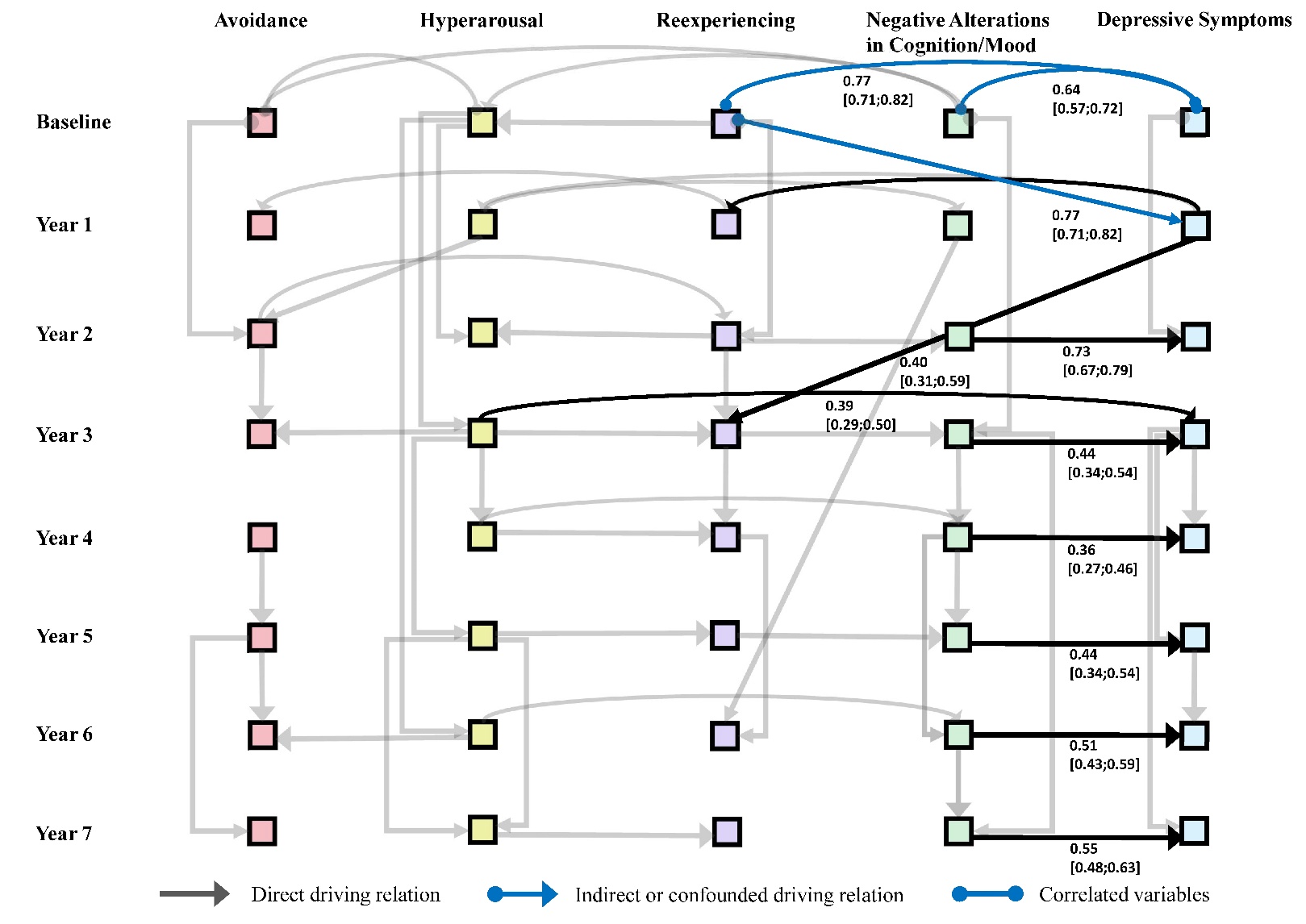

**Figure S7. Greedy Fast Causal Inference graph showing driving relations between NACM symptom cluster and depressive symptoms in the Mind Your Heart sample.** From years 2 through 7, negative alterations in cognition and mood (NACM) was the primary driver of depressive symptoms. The other DSM-V PTSD symptom clusters largely explained variance depressive symptoms through NACM. The rescaling of the PTSD checklist items from DSM-IV to DSM-V clusters is described in supplementary Figure S6. Paths connecting the DSM-V symptom clusters to depressive symptoms are bolded and presented with confidence intervals for the standardized path.

**Supplementary Code for “Causal discovery identifies posttraumatic stress as a driver of**

**internalizing symptoms across independent veteran and civilian populations”**

**Table of Contents**

R Code for Converting Tetrad DAG Output to Path Analysis…...………………………...	17

R Code for DSM-V Multilevel Confirmatory Factor Analysis on the PTSD Checklist…...	22

**R Code for Converting Tetrad DAG Output to Path Analysis**

setwd("C:/Users/pierc636/…") #WD HERE

Graph_Raw <- readLines("GRAPH.txt") #GRAPH FILE HERE (saved as .txt file)

Data <- read.csv("DATA.csv", head = T) #DATA FILE HERE

#install.packages("lavaan") #Do if needed

#install.packages("lavaanPlot") #Do if needed

#####THE REST OF THIS SHOULD RUN WITHOUT FURTHER USER INPUT#####

#####Will output excel files with "parameters" and "fit indices" in the working directory#####

#Importing graph edges as lavaan relations

Graph_Links <- Graph_Raw[5:(length(Graph_Raw)-4)]

Graph_Table <- as.data.frame(matrix(nrow = length(Graph_Links),

ncol = 10))

names(Graph_Table) <- c("Num","SVar","Rel","EVar",

paste("Extra",c(1:6), sep=""))

for(i in 1:length(Graph_Links)){

graph_bit <- unlist(strsplit(Graph_Links[i], split = " "))

Graph_Table[i,c(1:length(graph_bit))] <- graph_bit

}

Graph_Table$Rel[which(Graph_Table$Rel == "o->")] <- "~" #can also be "~~"#

Graph_Table$Rel[which(Graph_Table$Rel == "o-o")] <- "~~"

Graph_Table$Rel[which(Graph_Table$Rel == "-->")] <- "~"

#Making vector for lavaan

Graph_Lavaan <- rep(NA, times = (length(Graph_Links)))

#Filling vector

for(i in 1:nrow(Graph_Table)){

#flipping variable order from graph file - opposite sequence in r

Commas <- gsub(" ","",toString(Graph_Table[i,c(4,3,2)]))

Graph_Lavaan[i] <- gsub(",","",Commas)

}

####Run if excluding covariances not in graph####

#Making comparison vector so it knows what covariances to cancel out

AllVars <- unlist(strsplit(Graph_Raw[2],split=";"))

Graph_Covs <- as.data.frame(matrix(nrow =(length(AllVars)*length(AllVars)), ncol = 3))

for(i in 1:length(AllVars)){

if(i == 1){

Graph_Covs[c(1:length(AllVars)),1] <- AllVars[i]

Graph_Covs[c(1:length(AllVars)),2] <- "~~"

Graph_Covs[c(1:length(AllVars)),3] <- AllVars

} else {

Graph_Covs[c((1 + ((i-1)*length(AllVars))):(i*length(AllVars))),1] <- AllVars[i]

Graph_Covs[c((1 + ((i-1)*length(AllVars))):(i*length(AllVars))),2] <- "~~"

Graph_Covs[c((1 + ((i-1)*length(AllVars))):(i*length(AllVars))),3] <- AllVars

}}

#Making covariance vector to remove (relations left to right)

CoVars <- c(rep(NA, times = nrow(Graph_Covs)))

for(i in 1:length(CoVars)){

Covariance <- paste(Graph_Covs[i,1],Graph_Covs[i,2],Graph_Covs[i,3],sep="")

CoVars[i] <- Covariance

}

Covs_Matrix <- matrix(nrow=length(AllVars),ncol=length(AllVars))

for(i in 1:ncol(Covs_Matrix)){

if (i == 1){

Covs <- c(NA,CoVars[2:length(AllVars)])

Covs_Matrix[,i] <- Covs

} else {

if (i == ncol(Covs_Matrix)){

next

} else {

Covs <- CoVars[(((i-1)*length(AllVars))+1):(i*length(AllVars))]

CovSub <- c(rep(NA,times=i),Covs[(i+1):length(Covs)])

Covs_Matrix[,i] <- CovSub

}}}

#Making inverse covariance vector to remove (relations right to left)

I_CoVars <- c(rep(NA, times = nrow(Graph_Covs)))

for(i in 1:length(CoVars)){

I_Covariance <- paste(Graph_Covs[i,3],Graph_Covs[i,2],Graph_Covs[i,1],sep="")

I_CoVars[i] <- I_Covariance

}

I_Matrix <- matrix(nrow=length(AllVars),ncol=length(AllVars))

for(i in 1:ncol(I_Matrix)){

if (i == 1){

I_Covs <- c(NA,I_CoVars[2:length(AllVars)])

I_Matrix[,i] <- I_Covs

} else {

if (i == ncol(I_Matrix)){

next

} else {

I_Covs <- I_CoVars[(((i-1)*length(AllVars))+1):(i*length(AllVars))]

I_CovSub <- c(rep(NA,times=i),I_Covs[(i+1):length(I_Covs)])

I_Matrix[,i] <- I_CovSub

}}}

#Matrices to clean vectors

Covs_Vector <- as.vector(Covs_Matrix)

Covs_Vector <- Covs_Vector[which(is.na(Covs_Vector)==FALSE)]

I_Vector <- as.vector(I_Matrix)

I_Vector <- I_Vector[which(is.na(I_Vector)==FALSE)]

#Matching cov relations, identifying which to keep

Matches <- match(Covs_Vector,Graph_Lavaan)

I_Matches <- match(I_Vector,Graph_Lavaan)

for (i in 1:length(Matches)){

if(is.na(Matches[i])==TRUE){

Matches[i] <- I_Matches[i]

} else {

next

}}

#Taking out covariances specified in model from Covs_Vector

Vars_ToRm <- which(is.na(Matches)==TRUE)

Vars_Removing <- Covs_Vector[Vars_ToRm]

#Creating Final Lavaan Input

Main_Input <- paste(Graph_Lavaan,collapse = "\n")

Removal_Input <- gsub("~~","~~0*",toString(Vars_Removing))

Removal_Input <- gsub(", ","\n",Removal_Input)

Final_Input <- paste(Main_Input,"\n",Removal_Input, sep = "")

####if not excluding covariances not shown in graph, then just run this####

#Final_Input <- paste(Graph_Lavaan,collapse = "\n")

################################Running Model##############################

library(lavaan)

Clean_Model <- sem(Final_Input,data = Data,estimator = "MLR")

summary(Clean_Model, standardized=TRUE)

#Writing parameter and fit files

write.csv(standardizedSolution(Clean_Model),"Parameter_Estimates.csv")

write.csv(fitMeasures(Clean_Model),"Fit_Indices.csv")

#Plotting relations (very ugly plot)

library(lavaanPlot)

lavaanPlot(model = Clean_Model, node_options = list(shape = "box", fontname = "Helvetica"),

edge_options = list(color = "grey"), coefs = F)

**R Code for DSM-V Multilevel Confirmatory Factor Analysis on the PCL**

library(lavaan)

#Data = Mind Your Heart dataset

#Safety = Seeking Safety replication dataset

#PCL = PTSD checklist item

###DSMV Model in MYH Dataset###

M1 <- "

level:1

wP1 =~ 1*PCL1

wP2 =~ 1*PCL2

wP3 =~ 1*PCL3

wP4 =~ 1*PCL4

wP5 =~ 1*PCL5

wP6 =~ 1*PCL6

wP7 =~ 1*PCL7

wP8 =~ 1*PCL8

wP9 =~ 1*PCL9

wP10 =~ 1*PCL10

wP11 =~ 1*PCL11

wP12 =~ 1*PCL12

wP13 =~ 1*PCL13

wP14 =~ 1*PCL14

wP15 =~ 1*PCL15

wP16 =~ 1*PCL16

wP17 =~ 1*PCL17

PCL1~~0*PCL1

PCL2~~0*PCL2

PCL3~~0*PCL3

PCL4~~0*PCL4

PCL5~~0*PCL5

PCL6~~0*PCL6

PCL7~~0*PCL7

PCL8~~0*PCL8

PCL9~~0*PCL9

PCL10~~0*PCL10

PCL11~~0*PCL11

PCL12~~0*PCL12

PCL13~~0*PCL13

PCL14~~0*PCL14

PCL15~~0*PCL15

PCL16~~0*PCL16

PCL17~~0*PCL17

wR =~ wP1 + wP2 + wP3 + wP4 + wP5

wA =~ wP6 + wP7

wN =~ wP8 + wP9 + wP10 + wP11 + wP12

wH =~ wP13 + wP14 + wP15 + wP16 + wP17

level:2

bP1 =~ 1*PCL1

bP2 =~ 1*PCL2

bP3 =~ 1*PCL3

bP4 =~ 1*PCL4

bP5 =~ 1*PCL5

bP6 =~ 1*PCL6

bP7 =~ 1*PCL7

bP8 =~ 1*PCL8

bP9 =~ 1*PCL9

bP10 =~ 1*PCL10

bP11 =~ 1*PCL11

bP12 =~ 1*PCL12

bP13 =~ 1*PCL13

bP14 =~ 1*PCL14

bP15 =~ 1*PCL15

bP16 =~ 1*PCL16

bP17 =~ 1*PCL17

PCL1~~0*PCL1

PCL2~~0*PCL2

PCL3~~0*PCL3

PCL4~~0*PCL4

PCL5~~0*PCL5

PCL6~~0*PCL6

PCL7~~0*PCL7

PCL8~~0*PCL8

PCL9~~0*PCL9

PCL10~~0*PCL10

PCL11~~0*PCL11

PCL12~~0*PCL12

PCL13~~0*PCL13

PCL14~~0*PCL14

PCL15~~0*PCL15

PCL16~~0*PCL16

PCL17~~0*PCL17

bR =~ bP1 + bP2 + bP3 + bP4 + bP5

bA =~ bP6 + bP7

bN =~ bP8 + bP9 + bP10 + bP11 + bP12

bH =~ bP13 + bP14 + bP15 + bP16 + bP17"

fit_M1 <- cfa(M1, data=Data,cluster="ID",estimator="MLR")

fitMeasures(fit_M1)

mi_M1 <- modindices(fit_M1)

mi_M1[mi_M1$op == "=~",]

###DSMV model omitting problems remembering, concentration, sleep,

#and interest. Equalized loadings for all items except flashbacks. MYH Dataset.###

M2 <- "

level:1

wP1 =~ 1*PCL1

wP2 =~ 1*PCL2

wP3 =~ 1*PCL3

wP4 =~ 1*PCL4

wP5 =~ 1*PCL5

wP6 =~ 1*PCL6

wP7 =~ 1*PCL7

wP10 =~ 1*PCL10

wP11 =~ 1*PCL11

wP12 =~ 1*PCL12

wP14 =~ 1*PCL14

wP16 =~ 1*PCL16

wP17 =~ 1*PCL17

PCL1~~0*PCL1

PCL2~~0*PCL2

PCL3~~0*PCL3

PCL4~~0*PCL4

PCL5~~0*PCL5

PCL6~~0*PCL6

PCL7~~0*PCL7

PCL10~~0*PCL10

PCL11~~0*PCL11

PCL12~~0*PCL12

PCL14~~0*PCL14

PCL16~~0*PCL16

PCL17~~0*PCL17

wR =~ 1*wP1 + 1*wP2 + wP3 + 1*wP4 + 1*wP5

wA =~ 1*wP6 + 1*wP7

wN =~ 1*wP10 + 1*wP11 + 1*wP12

wH =~ 1*wP14 + 1*wP16 + 1*wP17

wP16~~wP17

level:2

bP1 =~ 1*PCL1

bP2 =~ 1*PCL2

bP3 =~ 1*PCL3

bP4 =~ 1*PCL4

bP5 =~ 1*PCL5

bP6 =~ 1*PCL6

bP7 =~ 1*PCL7

bP10 =~ 1*PCL10

bP11 =~ 1*PCL11

bP12 =~ 1*PCL12

bP14 =~ 1*PCL14

bP16 =~ 1*PCL16

bP17 =~ 1*PCL17

PCL1~~0*PCL1

PCL2~~0*PCL2

PCL3~~0*PCL3

PCL4~~0*PCL4

PCL5~~0*PCL5

PCL6~~0*PCL6

PCL7~~0*PCL7

PCL10~~0*PCL10

PCL11~~0*PCL11

PCL12~~0*PCL12

PCL14~~0*PCL14

PCL16~~0*PCL16

PCL17~~0*PCL17

bR =~ 1*bP1 + 1*bP2 + bP3 + 1*bP4 + 1*bP5

bA =~ 1*bP6 + 1*bP7

bN =~ 1*bP10 + 1*bP11 + 1*bP12

bH =~ 1*bP14 + 1*bP16 + 1*bP17

bP16~~bP17"

fit_M2 <- cfa(M2, data=Data,cluster="ID",estimator="MLR")

fitMeasures(fit_M2)

mi_M2 <- modindices(fit_M2)

mi_M2[mi_M2$op == "~~",]

###Replicating in WTTS Study###

MR <- "

level:1

wP1 =~ 1*PSS1

wP2 =~ 1*PSS2

wP3 =~ 1*PSS3

wP4 =~ 1*PSS4

wP5 =~ 1*PSS5

wP6 =~ 1*PSS6

wP7 =~ 1*PSS7

wP10 =~ 1*PSS10

wP11 =~ 1*PSS11

wP12 =~ 1*PSS12

wP14 =~ 1*PSS14

wP16 =~ 1*PSS16

wP17 =~ 1*PSS17

PSS1~~0*PSS1

PSS2~~0*PSS2

PSS3~~0*PSS3

PSS4~~0*PSS4

PSS5~~0*PSS5

PSS6~~0*PSS6

PSS7~~0*PSS7

PSS10~~0*PSS10

PSS11~~0*PSS11

PSS12~~0*PSS12

PSS14~~0*PSS14

PSS16~~0*PSS16

PSS17~~0*PSS17

wR =~ 1*wP1 + 1*wP2 + wP3 + 1*wP4 + 1*wP5

wA =~ 1*wP6 + 1*wP7

wN =~ 1*wP10 + 1*wP11 + 1*wP12

wH =~ 1*wP14 + 1*wP16 + 1*wP17

wP1~~wP4

wP16~~wP17

level:2

bP1 =~ 1*PSS1

bP2 =~ 1*PSS2

bP3 =~ 1*PSS3

bP4 =~ 1*PSS4

bP5 =~ 1*PSS5

bP6 =~ 1*PSS6

bP7 =~ 1*PSS7

bP10 =~ 1*PSS10

bP11 =~ 1*PSS11

bP12 =~ 1*PSS12

bP14 =~ 1*PSS14

bP16 =~ 1*PSS16

bP17 =~ 1*PSS17

PSS1~~0*PSS1

PSS2~~0*PSS2

PSS3~~0*PSS3

PSS4~~0*PSS4

PSS5~~0*PSS5

PSS6~~0*PSS6

PSS7~~0*PSS7

PSS10~~0*PSS10

PSS11~~0*PSS11

PSS12~~0*PSS12

PSS14~~0*PSS14

PSS16~~0*PSS16

PSS17~~0*PSS17

bR =~ 1*bP1 + 1*bP2 + bP3 + 1*bP4 + 1*bP5

bA =~ 1*bP6 + 1*bP7

bN =~ 1*bP10 + 1*bP11 + 1*bP12

bH =~ 1*bP14 + 1*bP16 + 1*bP17

bP1~~bP4

bP16~~bP17"

fit_MR <- cfa(MR, data=Safety,cluster="ID",estimator="MLR")

fitMeasures(fit_MR)

mi_MR <- modindices(fit_MR)

mi_MR[mi_MR$op == "~~",]

#Writing Data with DSM-V Rescoring#

Long <- Data

Long$R <- Long$PCL1 + Long$PCL2 + Long$PCL3 + Long$PCL4 + Long$PCL5

Long$A <- Long$PCL6 + Long$PCL7

Long$N <- Long$PCL10 + Long$PCL11 + Long$PCL12

Long$H <- Long$PCL14 + Long$PCL16 + Long$PCL17

Long$D <- Long$PHQ1 + Long$PHQ2 + Long$PHQ3 + Long$PHQ4 + Long$PHQ5 +

Long$PHQ6 + Long$PHQ7 + Long$PHQ8 + Long$PHQ9

Long_Rec <- Long[,c(1,2,29:33)]

Wide_Rec <- reshape(Long_Rec,timevar="Time",idvar="ID",direction="wide")

write.csv(Wide_Rec,"MYH_DSMV_Wide.csv",row.names=F)
