## Supplementary Table S2 for "Causal discovery identifies posttraumatic stress as a driver of internalizing symptoms across independent veteran and civilian populations"

Table S2. Stability analysis results on the MYH data

| Variable 1 | Variable 2 | <-- | --> | <-o | o-> | o-o | <-> | No Edge |
|---|---|---|---|---|---|---|---|---|
| Alcohol Use - BL | Alcohol Use - YR5 | 0.00 | 0.00 | 0.00 | 1.00 | 0.00 | 0.00 | 0.00 |
| Alcohol Use - BL | Alcohol Use - YR3 | 0.00 | 0.00 | 0.00 | 1.00 | 0.00 | 0.00 | 0.00 |
| Alcohol Use - BL | Alcohol Use - YR2 | 0.00 | 0.00 | 0.00 | 1.00 | 0.00 | 0.00 | 0.00 |
| Alcohol Use - BL | Alcohol Use - YR1 | 0.00 | 0.00 | 0.00 | 1.00 | 0.00 | 0.00 | 0.00 |
| Alcohol Use - BL | Alcohol Use - YR4 | 0.00 | 0.00 | 0.00 | 1.00 | 0.00 | 0.00 | 0.00 |
| Alcohol Use - BL | Alcohol Use - YR6 | 0.00 | 0.00 | 0.00 | 1.00 | 0.00 | 0.00 | 0.00 |
| Alcohol Use - BL | Alcohol Use - YR7 | 0.00 | 0.00 | 0.00 | 1.00 | 0.00 | 0.00 | 0.00 |
| PTSD Symptoms - YR1 | Depression - YR1 | 0.00 | 0.99 | 0.00 | 0.01 | 0.01 | 0.00 | 0.00 |
| Physical Amount - YR1 | Physical Compared - YR1 | 0.00 | 0.88 | 0.00 | 0.13 | 0.00 | 0.00 | 0.00 |
| PTSD Symptoms - YR2 | Depression - YR2 | 0.00 | 0.93 | 0.00 | 0.06 | 0.01 | 0.00 | 0.00 |
| PTSD Symptoms - YR3 | Depression - YR3 | 0.00 | 0.98 | 0.00 | 0.01 | 0.01 | 0.00 | 0.00 |
| PTSD Symptoms - YR5 | Depression - YR5 | 0.00 | 1.00 | 0.00 | 0.00 | 0.00 | 0.00 | 0.00 |
| Overall Health - BL | Quality of Life - BL | 0.00 | 0.00 | 0.01 | 0.00 | 0.99 | 0.00 | 0.00 |
| PTSD Symptoms - YR7 | Depression - YR7 | 0.01 | 0.99 | 0.00 | 0.00 | 0.00 | 0.00 | 0.00 |
| Physical Amount - YR2 | Physical Compared - YR2 | 0.00 | 0.89 | 0.00 | 0.11 | 0.00 | 0.00 | 0.00 |
| Overall Health - YR2 | Quality of Life - YR2 | 0.03 | 0.96 | 0.00 | 0.00 | 0.00 | 0.00 | 0.00 |
| Physical Limit - YR6 | Physical Limit - YR7 | 0.00 | 0.96 | 0.00 | 0.04 | 0.00 | 0.00 | 0.01 |
| Physical Amount - YR4 | Physical Compared - YR4 | 0.00 | 0.69 | 0.00 | 0.31 | 0.00 | 0.00 | 0.01 |
| Quality of Life - YR3 | Quality of Life - YR4 | 0.00 | 0.96 | 0.00 | 0.04 | 0.00 | 0.00 | 0.01 |
| Physical Limit - YR1 | Physical Limit - YR2 | 0.00 | 0.85 | 0.00 | 0.13 | 0.00 | 0.00 | 0.02 |
| PTSD Symptoms - BL | Depression - BL | 0.04 | 0.00 | 0.00 | 0.00 | 0.94 | 0.00 | 0.02 |
| PTSD Symptoms - YR1 | PTSD Symptoms - BL | 0.05 | 0.00 | 0.93 | 0.00 | 0.00 | 0.00 | 0.02 |
| PTSD Symptoms - YR6 | Depression - YR6 | 0.04 | 0.88 | 0.00 | 0.05 | 0.00 | 0.00 | 0.02 |
| Physical Compared - YR5 | Physical Compared - YR6 | 0.00 | 0.98 | 0.00 | 0.00 | 0.00 | 0.00 | 0.02 |
| Overall Health - YR5 | Quality of Life - YR5 | 0.24 | 0.70 | 0.00 | 0.03 | 0.00 | 0.00 | 0.03 |
| PTSD Symptoms - YR3 | PTSD Symptoms - YR4 | 0.00 | 0.92 | 0.00 | 0.02 | 0.00 | 0.00 | 0.06 |
| Physical Limit - YR1 | Physical Limit - YR3 | 0.00 | 0.80 | 0.00 | 0.13 | 0.00 | 0.00 | 0.07 |
| Overall Health - YR6 | Quality of Life - YR6 | 0.18 | 0.75 | 0.00 | 0.00 | 0.00 | 0.00 | 0.07 |
| Physical Limit - YR5 | Physical Limit - YR6 | 0.00 | 0.90 | 0.00 | 0.03 | 0.00 | 0.00 | 0.07 |
| PTSD Symptoms - YR6 | PTSD Symptoms - YR7 | 0.00 | 0.87 | 0.00 | 0.06 | 0.00 | 0.00 | 0.07 |
| Overall Health - YR3 | Physical Compared - YR3 | 0.00 | 0.90 | 0.00 | 0.03 | 0.00 | 0.00 | 0.07 |
| Physical Limit - YR5 | Physical Limit - YR4 | 0.70 | 0.00 | 0.21 | 0.00 | 0.00 | 0.00 | 0.09 |
| Overall Health - YR4 | Physical Compared - YR4 | 0.00 | 0.90 | 0.00 | 0.01 | 0.00 | 0.00 | 0.10 |
| Quality of Life - YR1 | Quality of Life - BL | 0.00 | 0.00 | 0.90 | 0.00 | 0.00 | 0.00 | 0.10 |
| Quality of Life - YR6 | Quality of Life - YR7 | 0.00 | 0.90 | 0.00 | 0.00 | 0.00 | 0.00 | 0.10 |
| PTSD Symptoms - BL | Social Function - YR4 | 0.00 | 0.05 | 0.00 | 0.85 | 0.00 | 0.00 | 0.10 |
| Physical Amount - YR3 | Physical Compared - YR3 | 0.00 | 0.58 | 0.00 | 0.31 | 0.00 | 0.00 | 0.10 |
| Overall Health - YR5 | Overall Health - YR6 | 0.00 | 0.86 | 0.00 | 0.04 | 0.00 | 0.00 | 0.11 |
| PTSD Symptoms - YR4 | PTSD Symptoms - YR5 | 0.00 | 0.66 | 0.00 | 0.22 | 0.00 | 0.00 | 0.12 |
| Overall Health - BL | Physical Limit - BL | 0.00 | 0.00 | 0.01 | 0.01 | 0.85 | 0.00 | 0.14 |
| Quality of Life - YR2 | Quality of Life - YR3 | 0.00 | 0.83 | 0.00 | 0.00 | 0.00 | 0.00 | 0.17 |
| Social Function - YR5 | PTSD Symptoms - BL | 0.04 | 0.00 | 0.78 | 0.00 | 0.00 | 0.00 | 0.18 |
| Physical Limit - YR1 | Physical Limit - BL | 0.01 | 0.00 | 0.81 | 0.00 | 0.00 | 0.00 | 0.18 |
| Social Function - YR3 | PTSD Symptoms - BL | 0.04 | 0.00 | 0.78 | 0.00 | 0.00 | 0.00 | 0.18 |
| Social Function - BL | Depression - BL | 0.04 | 0.00 | 0.00 | 0.00 | 0.78 | 0.00 | 0.18 |
| Overall Health - YR4 | Overall Health - YR3 | 0.78 | 0.00 | 0.03 | 0.00 | 0.00 | 0.00 | 0.19 |
| Overall Health - YR1 | Overall Health - BL | 0.01 | 0.00 | 0.80 | 0.00 | 0.00 | 0.00 | 0.19 |

Table S2. Stability analysis results on the MYH data

| Variable 1 | Variable 2 | <-- | --> | <-o | o-> | o-o | <-> | No Edge |
|---|---|---|---|---|---|---|---|---|
| Physical Amount - YR5 | Physical Compared - YR5 | 0.00 | 0.71 | 0.00 | 0.10 | 0.00 | 0.00 | 0.20 |
| Overall Health - YR6 | Overall Health - YR7 | 0.00 | 0.80 | 0.00 | 0.00 | 0.00 | 0.00 | 0.20 |
| Social Function - YR7 | PTSD Symptoms - BL | 0.04 | 0.00 | 0.76 | 0.00 | 0.00 | 0.00 | 0.21 |
| Overall Health - YR4 | Quality of Life - YR4 | 0.68 | 0.10 | 0.00 | 0.01 | 0.00 | 0.00 | 0.21 |
| Social Function - YR6 | PTSD Symptoms - BL | 0.04 | 0.00 | 0.75 | 0.00 | 0.00 | 0.00 | 0.21 |
| Overall Health - YR1 | Physical Compared - YR1 | 0.00 | 0.68 | 0.00 | 0.09 | 0.00 | 0.00 | 0.22 |
| Depression - YR3 | Depression - YR4 | 0.00 | 0.73 | 0.00 | 0.00 | 0.00 | 0.00 | 0.26 |
| Overall Health - YR2 | Physical Compared - YR2 | 0.00 | 0.73 | 0.00 | 0.00 | 0.00 | 0.00 | 0.27 |
| PTSD Symptoms - YR2 | PTSD Symptoms - YR3 | 0.00 | 0.63 | 0.00 | 0.10 | 0.00 | 0.00 | 0.27 |
| Depression - YR6 | Depression - YR7 | 0.00 | 0.71 | 0.00 | 0.00 | 0.00 | 0.01 | 0.28 |
| Overall Health - YR3 | Quality of Life - YR3 | 0.46 | 0.22 | 0.02 | 0.02 | 0.00 | 0.00 | 0.28 |
| Quality of Life - YR3 | Quality of Life - YR5 | 0.00 | 0.69 | 0.00 | 0.03 | 0.00 | 0.00 | 0.28 |
| Quality of Life - YR1 | Quality of Life - YR2 | 0.00 | 0.70 | 0.00 | 0.01 | 0.00 | 0.00 | 0.30 |
| Depression - YR4 | Social Function - YR4 | 0.69 | 0.00 | 0.00 | 0.00 | 0.00 | 0.00 | 0.31 |
| Physical Compared - YR5 | Physical Compared - YR7 | 0.00 | 0.69 | 0.00 | 0.00 | 0.00 | 0.00 | 0.31 |
| Physical Compared - YR3 | Physical Compared - YR5 | 0.00 | 0.68 | 0.00 | 0.00 | 0.00 | 0.00 | 0.32 |
| Physical Amount - YR1 | Physical Amount - YR2 | 0.00 | 0.60 | 0.00 | 0.08 | 0.00 | 0.00 | 0.32 |
| Overall Health - YR3 | Overall Health - BL | 0.02 | 0.00 | 0.62 | 0.00 | 0.00 | 0.00 | 0.36 |
| Quality of Life - BL | Depression - BL | 0.00 | 0.00 | 0.00 | 0.05 | 0.58 | 0.00 | 0.37 |
| Physical Limit - YR3 | Physical Limit - YR4 | 0.00 | 0.57 | 0.00 | 0.00 | 0.00 | 0.00 | 0.43 |
| Overall Health - YR2 | Overall Health - BL | 0.01 | 0.00 | 0.56 | 0.00 | 0.00 | 0.00 | 0.43 |
| Physical Amount - YR1 | Physical Limit - YR1 | 0.48 | 0.02 | 0.00 | 0.00 | 0.06 | 0.00 | 0.44 |
| Overall Health - YR2 | Physical Limit - YR2 | 0.43 | 0.13 | 0.00 | 0.00 | 0.00 | 0.00 | 0.44 |
| PTSD Symptoms - YR1 | PTSD Symptoms - YR2 | 0.00 | 0.48 | 0.00 | 0.06 | 0.00 | 0.00 | 0.46 |
| Overall Health - YR1 | Overall Health - YR5 | 0.00 | 0.46 | 0.00 | 0.07 | 0.00 | 0.00 | 0.47 |
| Depression - YR6 | Social Function - YR6 | 0.53 | 0.00 | 0.00 | 0.00 | 0.00 | 0.00 | 0.47 |
| Depression - YR4 | Quality of Life - YR4 | 0.04 | 0.48 | 0.00 | 0.00 | 0.00 | 0.00 | 0.48 |
| PTSD Symptoms - YR4 | PTSD Symptoms - YR7 | 0.00 | 0.50 | 0.00 | 0.01 | 0.00 | 0.00 | 0.49 |
| Physical Compared - YR5 | Physical Limit - YR5 | 0.11 | 0.39 | 0.00 | 0.00 | 0.00 | 0.00 | 0.50 |
| Overall Health - YR4 | Overall Health - YR6 | 0.00 | 0.50 | 0.00 | 0.00 | 0.00 | 0.00 | 0.50 |
| Physical Amount - YR1 | Physical Amount - YR4 | 0.00 | 0.43 | 0.00 | 0.07 | 0.00 | 0.00 | 0.50 |
| PTSD Symptoms - YR5 | PTSD Symptoms - YR6 | 0.00 | 0.48 | 0.00 | 0.01 | 0.00 | 0.00 | 0.51 |
| PTSD Symptoms - YR2 | PTSD Symptoms - BL | 0.06 | 0.00 | 0.43 | 0.00 | 0.00 | 0.00 | 0.51 |
| PTSD Symptoms - YR3 | Social Function - YR3 | 0.47 | 0.00 | 0.01 | 0.00 | 0.00 | 0.00 | 0.52 |
| Depression - YR2 | Social Function - YR2 | 0.39 | 0.00 | 0.09 | 0.00 | 0.00 | 0.00 | 0.53 |
| Overall Health - YR1 | Overall Health - YR2 | 0.00 | 0.38 | 0.00 | 0.09 | 0.00 | 0.00 | 0.53 |
| PTSD Symptoms - YR3 | PTSD Symptoms - YR5 | 0.00 | 0.44 | 0.00 | 0.01 | 0.00 | 0.00 | 0.56 |
| Physical Limit - YR3 | Physical Limit - YR5 | 0.00 | 0.44 | 0.00 | 0.00 | 0.00 | 0.00 | 0.56 |
| PTSD Symptoms - YR6 | Social Function - YR6 | 0.43 | 0.00 | 0.00 | 0.00 | 0.00 | 0.00 | 0.57 |
| Overall Health - YR3 | Overall Health - YR5 | 0.00 | 0.39 | 0.00 | 0.02 | 0.00 | 0.00 | 0.58 |
| Overall Health - YR1 | Quality of Life - YR1 | 0.25 | 0.13 | 0.00 | 0.03 | 0.01 | 0.00 | 0.59 |
| PTSD Symptoms - YR4 | Depression - YR4 | 0.04 | 0.36 | 0.00 | 0.01 | 0.00 | 0.00 | 0.59 |
| Physical Amount - YR1 | Quality of Life - YR1 | 0.37 | 0.02 | 0.00 | 0.01 | 0.00 | 0.00 | 0.60 |
| Physical Limit - YR3 | Social Function - YR3 | 0.38 | 0.00 | 0.02 | 0.00 | 0.00 | 0.00 | 0.61 |
| Quality of Life - YR3 | Quality of Life - YR6 | 0.00 | 0.36 | 0.00 | 0.01 | 0.00 | 0.00 | 0.62 |
| Overall Health - YR7 | Quality of Life - YR7 | 0.32 | 0.06 | 0.00 | 0.00 | 0.00 | 0.00 | 0.63 |
| PTSD Symptoms - YR1 | PTSD Symptoms - YR3 | 0.00 | 0.36 | 0.00 | 0.01 | 0.00 | 0.00 | 0.63 |

Table S2. Stability analysis results on the MYH data

| Variable 1 | Variable 2 | <-- | --> | <-o | o-> | o-o | <-> | No Edge |
|---|---|---|---|---|---|---|---|---|
| Physical Limit - YR1 | Physical Amount - YR7 | 0.00 | 0.33 | 0.00 | 0.04 | 0.00 | 0.00 | 0.63 |
| Physical Amount - YR1 | Physical Amount - YR6 | 0.00 | 0.32 | 0.00 | 0.05 | 0.00 | 0.00 | 0.63 |
| Social Function - YR2 | PTSD Symptoms - BL | 0.02 | 0.00 | 0.35 | 0.00 | 0.00 | 0.00 | 0.63 |
| PTSD Symptoms - YR1 | Social Function - YR1 | 0.13 | 0.00 | 0.23 | 0.00 | 0.00 | 0.00 | 0.64 |
| Overall Health - YR4 | Overall Health - YR2 | 0.36 | 0.00 | 0.00 | 0.00 | 0.00 | 0.00 | 0.64 |
| Quality of Life - YR5 | Quality of Life - YR6 | 0.00 | 0.35 | 0.00 | 0.00 | 0.00 | 0.00 | 0.65 |
| PTSD Symptoms - YR3 | PTSD Symptoms - YR6 | 0.00 | 0.33 | 0.00 | 0.01 | 0.00 | 0.00 | 0.66 |
| Depression - YR1 | Quality of Life - YR1 | 0.04 | 0.28 | 0.00 | 0.00 | 0.00 | 0.00 | 0.68 |
| Quality of Life - YR5 | Quality of Life - YR7 | 0.00 | 0.32 | 0.00 | 0.00 | 0.00 | 0.00 | 0.68 |
| Physical Limit - YR1 | Social Function - YR1 | 0.15 | 0.00 | 0.17 | 0.00 | 0.00 | 0.00 | 0.69 |
| Overall Health - YR1 | Physical Amount - YR5 | 0.00 | 0.27 | 0.00 | 0.05 | 0.00 | 0.00 | 0.69 |
| Physical Limit - YR6 | Physical Limit - YR4 | 0.29 | 0.00 | 0.02 | 0.00 | 0.00 | 0.00 | 0.70 |
| Overall Health - YR7 | Physical Compared - YR7 | 0.30 | 0.00 | 0.00 | 0.00 | 0.00 | 0.00 | 0.70 |
| Social Function - YR1 | PTSD Symptoms - BL | 0.01 | 0.00 | 0.28 | 0.00 | 0.00 | 0.00 | 0.71 |
| Physical Compared - YR3 | Physical Compared - YR7 | 0.00 | 0.29 | 0.00 | 0.00 | 0.00 | 0.00 | 0.71 |
| Quality of Life - YR3 | Quality of Life - YR7 | 0.00 | 0.28 | 0.00 | 0.01 | 0.00 | 0.00 | 0.71 |
| Depression - YR1 | Depression - BL | 0.02 | 0.00 | 0.27 | 0.00 | 0.00 | 0.00 | 0.72 |
| Quality of Life - YR3 | Quality of Life - BL | 0.00 | 0.00 | 0.28 | 0.00 | 0.00 | 0.00 | 0.72 |
| Physical Compared - YR7 | Physical Compared - YR4 | 0.28 | 0.00 | 0.00 | 0.00 | 0.00 | 0.00 | 0.72 |
| Social Function - YR2 | Social Function - BL | 0.02 | 0.00 | 0.26 | 0.00 | 0.00 | 0.00 | 0.73 |
| PTSD Symptoms - YR5 | Social Function - YR5 | 0.27 | 0.00 | 0.00 | 0.00 | 0.00 | 0.00 | 0.73 |
| Physical Compared - YR2 | Quality of Life - YR2 | 0.26 | 0.00 | 0.00 | 0.00 | 0.00 | 0.00 | 0.74 |
| Quality of Life - YR5 | Quality of Life - YR4 | 0.25 | 0.00 | 0.01 | 0.00 | 0.00 | 0.00 | 0.74 |
| Overall Health - YR4 | Overall Health - YR5 | 0.00 | 0.25 | 0.00 | 0.01 | 0.00 | 0.00 | 0.75 |
| Depression - YR2 | Depression - BL | 0.01 | 0.00 | 0.24 | 0.00 | 0.00 | 0.00 | 0.75 |
| Overall Health - YR2 | Overall Health - YR3 | 0.00 | 0.24 | 0.00 | 0.00 | 0.00 | 0.00 | 0.76 |
| Physical Compared - YR5 | Physical Compared - YR4 | 0.24 | 0.00 | 0.00 | 0.00 | 0.00 | 0.00 | 0.76 |
| PTSD Symptoms - BL | Social Function - BL | 0.00 | 0.01 | 0.00 | 0.00 | 0.23 | 0.00 | 0.76 |
| Physical Amount - YR3 | Overall Health - BL | 0.01 | 0.00 | 0.22 | 0.00 | 0.00 | 0.00 | 0.77 |
| Physical Limit - YR5 | Physical Limit - YR7 | 0.00 | 0.21 | 0.00 | 0.00 | 0.00 | 0.00 | 0.79 |
| Depression - YR7 | Social Function - YR7 | 0.20 | 0.00 | 0.00 | 0.00 | 0.00 | 0.00 | 0.80 |
| Physical Limit - YR3 | Physical Limit - BL | 0.01 | 0.00 | 0.19 | 0.00 | 0.00 | 0.00 | 0.80 |
| Overall Health - YR2 | Social Function - YR2 | 0.16 | 0.00 | 0.03 | 0.00 | 0.00 | 0.00 | 0.80 |
| Overall Health - YR1 | Overall Health - YR3 | 0.00 | 0.17 | 0.00 | 0.02 | 0.00 | 0.00 | 0.81 |
| PTSD Symptoms - YR2 | Social Function - YR2 | 0.18 | 0.00 | 0.01 | 0.00 | 0.00 | 0.00 | 0.81 |
| Physical Compared - YR3 | Physical Compared - YR6 | 0.00 | 0.19 | 0.00 | 0.00 | 0.00 | 0.00 | 0.81 |
| Social Function - YR7 | Depression - BL | 0.01 | 0.00 | 0.18 | 0.00 | 0.00 | 0.00 | 0.81 |
| PTSD Symptoms - YR1 | PTSD Symptoms - YR4 | 0.00 | 0.19 | 0.00 | 0.00 | 0.00 | 0.00 | 0.81 |
| Overall Health - YR3 | Physical Limit - YR3 | 0.06 | 0.13 | 0.00 | 0.00 | 0.00 | 0.00 | 0.81 |
| Depression - YR4 | Depression - YR5 | 0.00 | 0.19 | 0.00 | 0.00 | 0.00 | 0.00 | 0.81 |
| Physical Limit - YR6 | Social Function - YR6 | 0.19 | 0.00 | 0.00 | 0.00 | 0.00 | 0.00 | 0.81 |
| Overall Health - YR5 | Overall Health - YR7 | 0.00 | 0.18 | 0.00 | 0.01 | 0.00 | 0.00 | 0.81 |
| Depression - YR5 | Depression - YR6 | 0.00 | 0.18 | 0.00 | 0.00 | 0.00 | 0.00 | 0.82 |
| Depression - YR2 | Quality of Life - YR2 | 0.13 | 0.05 | 0.00 | 0.00 | 0.00 | 0.00 | 0.82 |
| Overall Health - YR3 | Overall Health - YR7 | 0.00 | 0.17 | 0.00 | 0.01 | 0.00 | 0.00 | 0.82 |
| Overall Health - YR2 | Depression - YR2 | 0.18 | 0.00 | 0.00 | 0.00 | 0.00 | 0.00 | 0.82 |
| Quality of Life - YR7 | Quality of Life - YR4 | 0.17 | 0.00 | 0.00 | 0.00 | 0.00 | 0.00 | 0.83 |

Table S2. Stability analysis results on the MYH data

| Variable 1 | Variable 2 | <-- | --> | <-o | o-> | o-o | <-> | No Edge |
|---|---|---|---|---|---|---|---|---|
| Quality of Life - YR2 | Depression - YR3 | 0.00 | 0.17 | 0.00 | 0.00 | 0.00 | 0.00 | 0.83 |
| Social Function - YR5 | Depression - BL | 0.01 | 0.00 | 0.17 | 0.00 | 0.00 | 0.00 | 0.83 |
| Depression - YR5 | Quality of Life - YR5 | 0.16 | 0.01 | 0.00 | 0.00 | 0.00 | 0.00 | 0.83 |
| Overall Health - YR5 | Overall Health - BL | 0.00 | 0.00 | 0.17 | 0.00 | 0.00 | 0.00 | 0.83 |
| Depression - YR3 | Depression - YR5 | 0.00 | 0.16 | 0.00 | 0.00 | 0.00 | 0.00 | 0.83 |
| Quality of Life - YR6 | Quality of Life - YR4 | 0.16 | 0.00 | 0.00 | 0.00 | 0.00 | 0.00 | 0.84 |
| Physical Amount - YR1 | Physical Amount - YR3 | 0.00 | 0.15 | 0.00 | 0.01 | 0.00 | 0.00 | 0.84 |
| Physical Limit - YR1 | Physical Amount - YR5 | 0.00 | 0.14 | 0.00 | 0.02 | 0.00 | 0.00 | 0.84 |
| Physical Limit - YR2 | Depression - YR2 | 0.11 | 0.04 | 0.00 | 0.00 | 0.00 | 0.00 | 0.84 |
| Social Function - YR6 | Depression - BL | 0.01 | 0.00 | 0.15 | 0.00 | 0.00 | 0.00 | 0.84 |
| Overall Health - YR4 | Overall Health - YR7 | 0.00 | 0.15 | 0.00 | 0.00 | 0.00 | 0.00 | 0.85 |
| Physical Limit - YR2 | Physical Limit - YR4 | 0.00 | 0.16 | 0.00 | 0.00 | 0.00 | 0.00 | 0.85 |
| Physical Limit - BL | Depression - BL | 0.00 | 0.00 | 0.00 | 0.05 | 0.11 | 0.00 | 0.85 |
| Physical Amount - YR5 | Physical Limit - BL | 0.00 | 0.00 | 0.15 | 0.00 | 0.00 | 0.00 | 0.85 |
| Quality of Life - YR1 | Quality of Life - YR3 | 0.00 | 0.14 | 0.00 | 0.00 | 0.00 | 0.00 | 0.85 |
| Quality of Life - YR2 | Social Function - YR2 | 0.11 | 0.00 | 0.03 | 0.00 | 0.00 | 0.00 | 0.85 |
| Physical Limit - YR2 | Social Function - YR2 | 0.11 | 0.00 | 0.04 | 0.00 | 0.00 | 0.00 | 0.86 |
| Depression - YR3 | Depression - BL | 0.00 | 0.00 | 0.14 | 0.00 | 0.00 | 0.00 | 0.86 |
| Physical Limit - YR4 | Social Function - YR4 | 0.14 | 0.00 | 0.00 | 0.00 | 0.00 | 0.00 | 0.86 |
| Depression - YR3 | Depression - YR6 | 0.00 | 0.14 | 0.00 | 0.00 | 0.00 | 0.00 | 0.86 |
| Social Function - YR1 | Physical Amount - YR7 | 0.00 | 0.06 | 0.00 | 0.08 | 0.00 | 0.00 | 0.86 |
| PTSD Symptoms - YR5 | PTSD Symptoms - YR7 | 0.00 | 0.14 | 0.00 | 0.00 | 0.00 | 0.00 | 0.86 |
| Physical Limit - YR2 | Physical Limit - BL | 0.01 | 0.00 | 0.12 | 0.00 | 0.00 | 0.00 | 0.87 |
| Quality of Life - YR2 | Quality of Life - YR7 | 0.00 | 0.13 | 0.00 | 0.00 | 0.00 | 0.00 | 0.87 |
| Depression - YR3 | Social Function - YR3 | 0.12 | 0.00 | 0.01 | 0.00 | 0.00 | 0.00 | 0.87 |
| PTSD Symptoms - YR4 | PTSD Symptoms - YR6 | 0.00 | 0.12 | 0.00 | 0.00 | 0.00 | 0.00 | 0.88 |
| Physical Limit - YR1 | Physical Amount - YR2 | 0.00 | 0.10 | 0.00 | 0.02 | 0.00 | 0.00 | 0.88 |
| Physical Compared - YR3 | Physical Limit - YR7 | 0.00 | 0.12 | 0.00 | 0.00 | 0.00 | 0.00 | 0.88 |
| Physical Compared - YR5 | Depression - YR5 | 0.00 | 0.11 | 0.00 | 0.00 | 0.00 | 0.00 | 0.89 |
| Overall Health - YR3 | Physical Compared - YR7 | 0.00 | 0.11 | 0.00 | 0.00 | 0.00 | 0.00 | 0.89 |
| Social Function - YR2 | Physical Limit - BL | 0.00 | 0.00 | 0.11 | 0.00 | 0.00 | 0.00 | 0.89 |
| Social Function - YR1 | Physical Limit - BL | 0.00 | 0.00 | 0.11 | 0.00 | 0.00 | 0.00 | 0.89 |
| Physical Amount - YR2 | Physical Limit - YR2 | 0.00 | 0.08 | 0.00 | 0.02 | 0.00 | 0.00 | 0.89 |
| Overall Health - YR1 | Physical Amount - YR3 | 0.00 | 0.09 | 0.00 | 0.02 | 0.00 | 0.00 | 0.90 |
| Physical Compared - YR6 | Physical Compared - YR4 | 0.10 | 0.00 | 0.00 | 0.00 | 0.00 | 0.00 | 0.90 |
| Physical Amount - YR1 | Physical Amount - YR7 | 0.00 | 0.09 | 0.00 | 0.01 | 0.00 | 0.00 | 0.90 |
| Physical Limit - YR7 | Social Function - YR7 | 0.10 | 0.00 | 0.00 | 0.00 | 0.00 | 0.00 | 0.90 |
| Physical Amount - YR3 | Quality of Life - BL | 0.00 | 0.00 | 0.10 | 0.00 | 0.00 | 0.00 | 0.90 |
| PTSD Symptoms - YR1 | Physical Limit - YR4 | 0.00 | 0.10 | 0.00 | 0.01 | 0.00 | 0.00 | 0.90 |
| PTSD Symptoms - YR3 | PTSD Symptoms - BL | 0.01 | 0.00 | 0.09 | 0.00 | 0.00 | 0.00 | 0.90 |
| Quality of Life - YR1 | Physical Amount - YR7 | 0.00 | 0.10 | 0.00 | 0.00 | 0.00 | 0.00 | 0.90 |
| PTSD Symptoms - YR2 | PTSD Symptoms - YR4 | 0.00 | 0.09 | 0.00 | 0.01 | 0.00 | 0.00 | 0.90 |
| Depression - YR6 | Quality of Life - YR6 | 0.00 | 0.10 | 0.00 | 0.00 | 0.00 | 0.00 | 0.90 |
| PTSD Symptoms - BL | Quality of Life - BL | 0.00 | 0.00 | 0.00 | 0.00 | 0.10 | 0.00 | 0.90 |
| Depression - YR5 | Social Function - YR5 | 0.10 | 0.00 | 0.00 | 0.00 | 0.00 | 0.00 | 0.90 |
| PTSD Symptoms - YR3 | Depression - YR4 | 0.00 | 0.09 | 0.00 | 0.00 | 0.00 | 0.00 | 0.91 |
| Social Function - YR1 | Depression - BL | 0.01 | 0.00 | 0.09 | 0.00 | 0.00 | 0.00 | 0.91 |

Table S2. Stability analysis results on the MYH data

| Variable 1 | Variable 2 | <-- | --> | <-o | o-> | o-o | <-> | No Edge |
|---|---|---|---|---|---|---|---|---|
| Physical Limit - YR1 | Physical Amount - YR4 | 0.00 | 0.08 | 0.00 | 0.01 | 0.00 | 0.00 | 0.91 |
| Overall Health - YR6 | Physical Compared - YR6 | 0.09 | 0.00 | 0.00 | 0.00 | 0.00 | 0.00 | 0.91 |
| Overall Health - BL | PTSD Symptoms - BL | 0.00 | 0.00 | 0.00 | 0.00 | 0.09 | 0.00 | 0.91 |
| Overall Health - YR2 | Physical Limit - YR3 | 0.00 | 0.08 | 0.00 | 0.00 | 0.00 | 0.00 | 0.92 |
| Social Function - YR1 | Physical Amount - YR5 | 0.00 | 0.04 | 0.00 | 0.04 | 0.00 | 0.00 | 0.92 |
| PTSD Symptoms - YR6 | Quality of Life - YR6 | 0.04 | 0.04 | 0.00 | 0.00 | 0.00 | 0.00 | 0.92 |
| Physical Limit - YR3 | Physical Limit - YR6 | 0.00 | 0.08 | 0.00 | 0.00 | 0.00 | 0.00 | 0.92 |
| Overall Health - BL | Social Function - BL | 0.00 | 0.00 | 0.00 | 0.00 | 0.08 | 0.00 | 0.92 |
| PTSD Symptoms - YR6 | Physical Limit - YR6 | 0.01 | 0.06 | 0.00 | 0.01 | 0.00 | 0.00 | 0.93 |
| Physical Amount - YR5 | Quality of Life - BL | 0.00 | 0.00 | 0.07 | 0.00 | 0.00 | 0.00 | 0.93 |
| Physical Limit - YR6 | Depression - YR6 | 0.07 | 0.00 | 0.00 | 0.00 | 0.00 | 0.00 | 0.93 |
| Depression - BL | Social Function - YR4 | 0.00 | 0.00 | 0.00 | 0.07 | 0.00 | 0.00 | 0.93 |
| Physical Limit - YR1 | Physical Limit - YR4 | 0.00 | 0.06 | 0.00 | 0.01 | 0.00 | 0.00 | 0.93 |
| Depression - YR2 | Depression - YR4 | 0.00 | 0.07 | 0.00 | 0.00 | 0.00 | 0.00 | 0.93 |
| Depression - YR2 | PTSD Symptoms - YR7 | 0.00 | 0.07 | 0.00 | 0.00 | 0.00 | 0.00 | 0.93 |
| PTSD Symptoms - BL | Physical Limit - BL | 0.00 | 0.00 | 0.00 | 0.00 | 0.06 | 0.00 | 0.93 |
| Physical Limit - YR2 | Physical Limit - YR3 | 0.00 | 0.06 | 0.00 | 0.01 | 0.00 | 0.00 | 0.93 |
| Quality of Life - YR2 | Quality of Life - YR5 | 0.00 | 0.07 | 0.00 | 0.00 | 0.00 | 0.00 | 0.93 |
| Overall Health - YR3 | Overall Health - YR6 | 0.00 | 0.06 | 0.00 | 0.00 | 0.00 | 0.00 | 0.94 |
| Quality of Life - YR2 | Quality of Life - BL | 0.00 | 0.00 | 0.06 | 0.00 | 0.00 | 0.00 | 0.94 |
| Social Function - YR2 | Physical Limit - YR4 | 0.00 | 0.06 | 0.00 | 0.01 | 0.00 | 0.00 | 0.94 |
| Social Function - YR3 | Physical Limit - YR4 | 0.00 | 0.06 | 0.00 | 0.01 | 0.00 | 0.00 | 0.94 |
| PTSD Symptoms - YR1 | PTSD Symptoms - YR7 | 0.00 | 0.06 | 0.00 | 0.00 | 0.00 | 0.00 | 0.94 |
| Quality of Life - YR2 | Depression - YR5 | 0.00 | 0.06 | 0.00 | 0.00 | 0.00 | 0.00 | 0.94 |
| Social Function - YR3 | Social Function - BL | 0.00 | 0.00 | 0.06 | 0.00 | 0.00 | 0.00 | 0.94 |
| Overall Health - YR4 | Overall Health - YR1 | 0.04 | 0.00 | 0.01 | 0.00 | 0.00 | 0.00 | 0.94 |
| Overall Health - YR6 | Social Function - YR6 | 0.05 | 0.00 | 0.00 | 0.00 | 0.00 | 0.00 | 0.95 |
| Overall Health - YR2 | Physical Compared - YR3 | 0.00 | 0.05 | 0.00 | 0.00 | 0.00 | 0.00 | 0.95 |
| Physical Compared - YR2 | Physical Compared - YR5 | 0.00 | 0.05 | 0.00 | 0.00 | 0.00 | 0.00 | 0.95 |
| PTSD Symptoms - YR7 | PTSD Symptoms - BL | 0.00 | 0.00 | 0.05 | 0.00 | 0.00 | 0.00 | 0.95 |
| Physical Limit - YR5 | Physical Limit - BL | 0.00 | 0.00 | 0.05 | 0.00 | 0.00 | 0.00 | 0.95 |
| Quality of Life - YR2 | Quality of Life - YR4 | 0.00 | 0.05 | 0.00 | 0.00 | 0.00 | 0.00 | 0.95 |
| Depression - YR1 | Physical Amount - YR4 | 0.00 | 0.05 | 0.00 | 0.00 | 0.00 | 0.00 | 0.95 |
| Overall Health - YR3 | Physical Compared - YR4 | 0.00 | 0.05 | 0.00 | 0.00 | 0.00 | 0.00 | 0.95 |
| PTSD Symptoms - YR5 | PTSD Symptoms - BL | 0.00 | 0.00 | 0.05 | 0.00 | 0.00 | 0.00 | 0.95 |
| Overall Health - YR6 | Physical Limit - YR4 | 0.04 | 0.00 | 0.00 | 0.00 | 0.00 | 0.00 | 0.95 |
| Overall Health - YR1 | Physical Amount - YR2 | 0.00 | 0.04 | 0.00 | 0.00 | 0.00 | 0.00 | 0.96 |
| Quality of Life - YR1 | Physical Amount - YR6 | 0.00 | 0.04 | 0.00 | 0.00 | 0.00 | 0.00 | 0.96 |
| Social Function - YR3 | Depression - BL | 0.00 | 0.00 | 0.04 | 0.00 | 0.00 | 0.00 | 0.96 |
| Physical Amount - YR6 | Physical Limit - YR6 | 0.00 | 0.02 | 0.00 | 0.02 | 0.00 | 0.00 | 0.96 |
| Social Function - YR2 | Depression - BL | 0.00 | 0.00 | 0.04 | 0.00 | 0.00 | 0.00 | 0.96 |
| Overall Health - YR3 | Depression - YR3 | 0.03 | 0.01 | 0.00 | 0.00 | 0.00 | 0.00 | 0.96 |
| Overall Health - BL | Depression - BL | 0.00 | 0.00 | 0.00 | 0.00 | 0.04 | 0.00 | 0.96 |
| Physical Limit - YR3 | Depression - YR3 | 0.04 | 0.00 | 0.00 | 0.00 | 0.00 | 0.00 | 0.96 |
| Depression - YR5 | Depression - YR7 | 0.00 | 0.04 | 0.00 | 0.00 | 0.00 | 0.00 | 0.96 |
| Depression - YR5 | Depression - BL | 0.00 | 0.00 | 0.04 | 0.00 | 0.00 | 0.00 | 0.96 |
| Physical Amount - YR1 | Physical Amount - YR5 | 0.00 | 0.04 | 0.00 | 0.00 | 0.00 | 0.00 | 0.96 |

Table S2. Stability analysis results on the MYH data

| Variable 1 | Variable 2 | <-- | --> | <-o | o-> | o-o | <-> | No Edge |
|---|---|---|---|---|---|---|---|---|
| Overall Health - YR2 | Overall Health - YR6 | 0.00 | 0.04 | 0.00 | 0.00 | 0.00 | 0.00 | 0.96 |
| Overall Health - YR5 | Physical Compared - YR6 | 0.00 | 0.04 | 0.00 | 0.00 | 0.00 | 0.00 | 0.96 |
| Depression - YR3 | Quality of Life - YR3 | 0.04 | 0.00 | 0.00 | 0.00 | 0.00 | 0.00 | 0.96 |
| Physical Amount - YR7 | Overall Health - BL | 0.00 | 0.00 | 0.04 | 0.00 | 0.00 | 0.00 | 0.96 |
| Depression - YR1 | Physical Amount - YR7 | 0.00 | 0.04 | 0.00 | 0.00 | 0.00 | 0.00 | 0.96 |
| Physical Compared - YR3 | Physical Limit - YR3 | 0.04 | 0.00 | 0.00 | 0.00 | 0.00 | 0.00 | 0.96 |
| Physical Compared - YR6 | Quality of Life - YR6 | 0.00 | 0.04 | 0.00 | 0.00 | 0.00 | 0.00 | 0.96 |
| PTSD Symptoms - YR1 | Physical Amount - YR4 | 0.00 | 0.04 | 0.00 | 0.00 | 0.00 | 0.00 | 0.96 |
| Physical Amount - YR7 | Quality of Life - BL | 0.00 | 0.00 | 0.04 | 0.00 | 0.00 | 0.00 | 0.96 |
| Physical Limit - YR1 | Quality of Life - YR1 | 0.04 | 0.00 | 0.00 | 0.00 | 0.00 | 0.00 | 0.96 |
| Physical Limit - YR2 | Physical Limit - YR6 | 0.00 | 0.04 | 0.00 | 0.00 | 0.00 | 0.00 | 0.96 |
| Overall Health - YR2 | Depression - YR3 | 0.00 | 0.04 | 0.00 | 0.00 | 0.00 | 0.00 | 0.96 |
| Quality of Life - YR5 | Social Function - YR5 | 0.04 | 0.00 | 0.00 | 0.00 | 0.00 | 0.00 | 0.96 |
| Overall Health - YR1 | Physical Limit - YR1 | 0.01 | 0.02 | 0.00 | 0.01 | 0.00 | 0.00 | 0.97 |
| Overall Health - YR2 | Overall Health - YR5 | 0.00 | 0.03 | 0.00 | 0.00 | 0.00 | 0.00 | 0.97 |
| Physical Limit - YR2 | Physical Limit - YR5 | 0.00 | 0.03 | 0.00 | 0.00 | 0.00 | 0.00 | 0.97 |
| Physical Compared - YR7 | Physical Limit - YR7 | 0.00 | 0.03 | 0.00 | 0.00 | 0.00 | 0.00 | 0.97 |
| Physical Amount - YR4 | Physical Limit - YR4 | 0.00 | 0.02 | 0.00 | 0.01 | 0.00 | 0.00 | 0.97 |
| Physical Amount - YR5 | Overall Health - BL | 0.00 | 0.00 | 0.03 | 0.00 | 0.00 | 0.00 | 0.97 |
| Quality of Life - YR1 | Social Function - YR1 | 0.01 | 0.00 | 0.02 | 0.00 | 0.00 | 0.00 | 0.97 |
| Overall Health - YR6 | PTSD Symptoms - YR6 | 0.02 | 0.01 | 0.00 | 0.00 | 0.00 | 0.00 | 0.97 |
| Physical Amount - YR6 | Physical Limit - BL | 0.00 | 0.00 | 0.03 | 0.00 | 0.00 | 0.00 | 0.97 |
| Physical Compared - YR1 | Physical Limit - YR1 | 0.02 | 0.00 | 0.01 | 0.00 | 0.00 | 0.00 | 0.97 |
| Physical Amount - YR2 | Physical Compared - YR3 | 0.00 | 0.03 | 0.00 | 0.01 | 0.00 | 0.00 | 0.97 |
| Physical Amount - YR3 | Physical Limit - BL | 0.00 | 0.00 | 0.03 | 0.00 | 0.00 | 0.00 | 0.97 |
| Physical Compared - YR1 | Physical Compared - YR7 | 0.00 | 0.03 | 0.00 | 0.00 | 0.00 | 0.00 | 0.97 |
| Overall Health - YR5 | Physical Compared - YR5 | 0.01 | 0.02 | 0.00 | 0.00 | 0.00 | 0.00 | 0.97 |
| Physical Limit - YR3 | Physical Limit - YR7 | 0.00 | 0.03 | 0.00 | 0.00 | 0.00 | 0.00 | 0.97 |
| Overall Health - YR7 | Overall Health - BL | 0.00 | 0.00 | 0.03 | 0.00 | 0.00 | 0.00 | 0.97 |
| Social Function - BL | Social Function - YR4 | 0.00 | 0.00 | 0.00 | 0.03 | 0.00 | 0.00 | 0.97 |
| Physical Limit - YR7 | Depression - YR7 | 0.03 | 0.00 | 0.00 | 0.00 | 0.00 | 0.00 | 0.97 |
| Overall Health - YR1 | Overall Health - YR7 | 0.00 | 0.03 | 0.00 | 0.00 | 0.00 | 0.00 | 0.97 |
| Physical Compared - YR5 | Quality of Life - YR5 | 0.01 | 0.02 | 0.00 | 0.00 | 0.00 | 0.00 | 0.97 |
| Overall Health - YR7 | Physical Limit - YR7 | 0.01 | 0.02 | 0.00 | 0.00 | 0.00 | 0.00 | 0.97 |
| Overall Health - YR3 | Social Function - YR3 | 0.03 | 0.00 | 0.00 | 0.00 | 0.00 | 0.00 | 0.98 |
| PTSD Symptoms - YR4 | PTSD Symptoms - BL | 0.00 | 0.00 | 0.02 | 0.00 | 0.00 | 0.00 | 0.98 |
| Alcohol Use - YR1 | Social Function - BL | 0.00 | 0.00 | 0.02 | 0.00 | 0.00 | 0.00 | 0.98 |
| Overall Health - YR2 | Depression - YR5 | 0.00 | 0.02 | 0.00 | 0.00 | 0.00 | 0.00 | 0.98 |
| Overall Health - YR4 | Physical Limit - YR4 | 0.01 | 0.01 | 0.00 | 0.00 | 0.00 | 0.00 | 0.98 |
| Quality of Life - YR7 | Quality of Life - BL | 0.00 | 0.00 | 0.02 | 0.00 | 0.00 | 0.00 | 0.98 |
| Quality of Life - BL | Social Function - BL | 0.00 | 0.00 | 0.00 | 0.00 | 0.02 | 0.00 | 0.98 |
| Quality of Life - YR1 | Quality of Life - YR7 | 0.00 | 0.02 | 0.00 | 0.00 | 0.00 | 0.00 | 0.98 |
| PTSD Symptoms - YR6 | PTSD Symptoms - BL | 0.00 | 0.00 | 0.02 | 0.00 | 0.00 | 0.00 | 0.98 |
| PTSD Symptoms - YR7 | Physical Limit - YR7 | 0.02 | 0.00 | 0.00 | 0.00 | 0.00 | 0.00 | 0.98 |
| Overall Health - YR4 | Quality of Life - YR5 | 0.00 | 0.02 | 0.00 | 0.00 | 0.00 | 0.00 | 0.98 |
| Overall Health - YR4 | Overall Health - BL | 0.00 | 0.00 | 0.02 | 0.00 | 0.00 | 0.00 | 0.98 |
| Physical Compared - YR1 | Overall Health - BL | 0.00 | 0.00 | 0.02 | 0.00 | 0.00 | 0.00 | 0.98 |

Table S2. Stability analysis results on the MYH data

| Variable 1 | Variable 2 | <-- | --> | <-o | o-> | o-o | <-> | No Edge |
|---|---|---|---|---|---|---|---|---|
| Social Function - YR6 | Quality of Life - BL | 0.00 | 0.00 | 0.02 | 0.00 | 0.00 | 0.00 | 0.98 |
| Physical Limit - YR1 | Depression - YR1 | 0.02 | 0.00 | 0.00 | 0.00 | 0.00 | 0.00 | 0.98 |
| Physical Limit - YR1 | Physical Limit - YR5 | 0.00 | 0.02 | 0.00 | 0.00 | 0.00 | 0.00 | 0.98 |
| Quality of Life - YR1 | Physical Compared - YR4 | 0.00 | 0.02 | 0.00 | 0.00 | 0.00 | 0.00 | 0.98 |
| Physical Amount - YR2 | Depression - BL | 0.00 | 0.00 | 0.02 | 0.00 | 0.00 | 0.00 | 0.98 |
| Depression - YR2 | Depression - YR3 | 0.00 | 0.02 | 0.00 | 0.00 | 0.00 | 0.00 | 0.98 |
| Overall Health - YR3 | Depression - YR5 | 0.00 | 0.02 | 0.00 | 0.00 | 0.00 | 0.00 | 0.98 |
| PTSD Symptoms - YR3 | PTSD Symptoms - YR7 | 0.00 | 0.02 | 0.00 | 0.00 | 0.00 | 0.00 | 0.98 |
| PTSD Symptoms - YR5 | Physical Limit - YR5 | 0.02 | 0.00 | 0.00 | 0.00 | 0.00 | 0.00 | 0.98 |
| Physical Limit - YR5 | Social Function - YR5 | 0.02 | 0.00 | 0.00 | 0.00 | 0.00 | 0.00 | 0.98 |
| Physical Limit - YR2 | Quality of Life - YR2 | 0.02 | 0.00 | 0.00 | 0.00 | 0.00 | 0.00 | 0.98 |
| Physical Limit - YR1 | Physical Amount - YR6 | 0.00 | 0.02 | 0.00 | 0.00 | 0.00 | 0.00 | 0.98 |
| Quality of Life - YR1 | Quality of Life - YR4 | 0.00 | 0.02 | 0.00 | 0.00 | 0.00 | 0.00 | 0.98 |
| Social Function - YR1 | Social Function - BL | 0.00 | 0.00 | 0.02 | 0.00 | 0.00 | 0.00 | 0.98 |
| PTSD Symptoms - YR2 | PTSD Symptoms - YR7 | 0.00 | 0.02 | 0.00 | 0.00 | 0.00 | 0.00 | 0.98 |
| Depression - YR6 | Physical Limit - YR7 | 0.00 | 0.02 | 0.00 | 0.00 | 0.00 | 0.00 | 0.98 |
| Social Function - YR6 | Social Function - BL | 0.00 | 0.00 | 0.02 | 0.00 | 0.00 | 0.00 | 0.98 |
| Overall Health - YR1 | Quality of Life - BL | 0.00 | 0.00 | 0.02 | 0.00 | 0.00 | 0.00 | 0.98 |
| Physical Amount - YR1 | Depression - YR1 | 0.01 | 0.00 | 0.00 | 0.00 | 0.00 | 0.00 | 0.98 |
| Social Function - YR1 | Physical Amount - YR6 | 0.00 | 0.01 | 0.00 | 0.00 | 0.00 | 0.00 | 0.98 |
| Social Function - YR3 | Physical Limit - BL | 0.00 | 0.00 | 0.02 | 0.00 | 0.00 | 0.00 | 0.98 |
| Physical Compared - YR1 | Quality of Life - YR1 | 0.01 | 0.00 | 0.00 | 0.00 | 0.00 | 0.00 | 0.99 |
| Social Function - YR3 | Quality of Life - YR4 | 0.00 | 0.01 | 0.00 | 0.00 | 0.00 | 0.00 | 0.99 |
| Physical Amount - YR5 | Physical Limit - YR5 | 0.00 | 0.01 | 0.00 | 0.00 | 0.00 | 0.00 | 0.99 |
| Depression - YR5 | Quality of Life - YR7 | 0.00 | 0.01 | 0.00 | 0.00 | 0.00 | 0.00 | 0.99 |
| Social Function - YR6 | Overall Health - BL | 0.00 | 0.00 | 0.01 | 0.00 | 0.00 | 0.00 | 0.99 |
| Physical Amount - YR7 | Depression - BL | 0.00 | 0.00 | 0.01 | 0.00 | 0.00 | 0.00 | 0.99 |
| Physical Limit - BL | Social Function - BL | 0.00 | 0.00 | 0.00 | 0.00 | 0.01 | 0.00 | 0.99 |
| Overall Health - YR1 | Physical Compared - YR4 | 0.00 | 0.01 | 0.00 | 0.00 | 0.00 | 0.00 | 0.99 |
| Physical Amount - YR1 | Physical Limit - BL | 0.00 | 0.00 | 0.01 | 0.00 | 0.00 | 0.00 | 0.99 |
| Social Function - YR1 | PTSD Symptoms - YR2 | 0.00 | 0.00 | 0.00 | 0.01 | 0.00 | 0.00 | 0.99 |
| Physical Compared - YR3 | Depression - YR5 | 0.00 | 0.01 | 0.00 | 0.00 | 0.00 | 0.00 | 0.99 |
| Overall Health - YR1 | Quality of Life - YR7 | 0.00 | 0.01 | 0.00 | 0.00 | 0.00 | 0.00 | 0.99 |
| Physical Limit - YR1 | Physical Amount - YR3 | 0.00 | 0.01 | 0.00 | 0.00 | 0.00 | 0.00 | 0.99 |
| Depression - YR1 | PTSD Symptoms - YR4 | 0.00 | 0.01 | 0.00 | 0.00 | 0.00 | 0.00 | 0.99 |
| Physical Amount - YR2 | Overall Health - BL | 0.00 | 0.00 | 0.01 | 0.00 | 0.00 | 0.00 | 0.99 |
| Physical Amount - YR5 | PTSD Symptoms - BL | 0.00 | 0.00 | 0.01 | 0.00 | 0.00 | 0.00 | 0.99 |
| Physical Amount - YR5 | Depression - BL | 0.00 | 0.00 | 0.01 | 0.00 | 0.00 | 0.00 | 0.99 |
| Overall Health - YR6 | Physical Limit - YR6 | 0.01 | 0.00 | 0.00 | 0.00 | 0.00 | 0.00 | 0.99 |
| Overall Health - YR4 | Physical Limit - YR7 | 0.00 | 0.01 | 0.00 | 0.00 | 0.00 | 0.00 | 0.99 |
| Physical Limit - YR1 | Physical Limit - YR6 | 0.00 | 0.01 | 0.00 | 0.00 | 0.00 | 0.00 | 0.99 |
| Social Function - YR1 | Quality of Life - BL | 0.00 | 0.00 | 0.01 | 0.00 | 0.00 | 0.00 | 0.99 |
| Overall Health - YR3 | Quality of Life - YR6 | 0.00 | 0.01 | 0.00 | 0.00 | 0.00 | 0.00 | 0.99 |
| Physical Amount - YR5 | Physical Compared - YR6 | 0.00 | 0.01 | 0.00 | 0.00 | 0.00 | 0.00 | 0.99 |
| PTSD Symptoms - YR7 | Social Function - YR7 | 0.01 | 0.00 | 0.00 | 0.00 | 0.00 | 0.00 | 0.99 |
| Social Function - BL | Physical Limit - YR4 | 0.00 | 0.00 | 0.00 | 0.01 | 0.00 | 0.00 | 0.99 |
| Physical Limit - YR2 | Depression - BL | 0.00 | 0.00 | 0.01 | 0.00 | 0.00 | 0.00 | 0.99 |

Table S3. Stability analysis results on the MYH data

| Variable 1 | Variable 2 | <-- | --> | <-o | o-> | o-o | <-> | No Edge |
|---|---|---|---|---|---|---|---|---|
| Physical Amount - YR3 | Physical Limit - YR3 | 0.00 | 0.01 | 0.00 | 0.00 | 0.00 | 0.00 | 0.99 |
| PTSD Symptoms - YR4 | Physical Limit - YR4 | 0.00 | 0.01 | 0.00 | 0.00 | 0.00 | 0.00 | 0.99 |
| PTSD Symptoms - YR5 | Social Function - BL | 0.00 | 0.00 | 0.01 | 0.00 | 0.00 | 0.00 | 0.99 |
| Physical Amount - YR5 | Physical Compared - YR7 | 0.00 | 0.01 | 0.00 | 0.00 | 0.00 | 0.00 | 0.99 |
| Physical Amount - YR5 | Quality of Life - YR5 | 0.00 | 0.01 | 0.00 | 0.00 | 0.00 | 0.00 | 0.99 |
| Quality of Life - YR6 | Social Function - YR6 | 0.01 | 0.00 | 0.00 | 0.00 | 0.00 | 0.00 | 0.99 |
| Alcohol Use - BL | PTSD Symptoms - BL | 0.00 | 0.00 | 0.00 | 0.00 | 0.01 | 0.00 | 0.99 |
| PTSD Symptoms - YR1 | PTSD Symptoms - YR6 | 0.00 | 0.01 | 0.00 | 0.00 | 0.00 | 0.00 | 0.99 |
| Social Function - YR1 | Physical Limit - YR4 | 0.00 | 0.00 | 0.00 | 0.01 | 0.00 | 0.00 | 0.99 |
| Overall Health - YR2 | PTSD Symptoms - YR2 | 0.01 | 0.00 | 0.00 | 0.00 | 0.00 | 0.00 | 0.99 |
| PTSD Symptoms - YR2 | PTSD Symptoms - YR6 | 0.00 | 0.01 | 0.00 | 0.00 | 0.00 | 0.00 | 0.99 |
| Physical Limit - YR2 | Physical Limit - YR7 | 0.00 | 0.01 | 0.00 | 0.00 | 0.00 | 0.00 | 0.99 |
| PTSD Symptoms - YR4 | Quality of Life - YR4 | 0.00 | 0.01 | 0.00 | 0.00 | 0.00 | 0.00 | 0.99 |
| Physical Limit - YR6 | PTSD Symptoms - YR7 | 0.00 | 0.01 | 0.00 | 0.00 | 0.00 | 0.00 | 0.99 |
| Social Function - BL | Physical Amount - YR4 | 0.00 | 0.00 | 0.00 | 0.01 | 0.00 | 0.00 | 0.99 |
| PTSD Symptoms - YR1 | Physical Limit - YR1 | 0.00 | 0.01 | 0.00 | 0.00 | 0.00 | 0.00 | 0.99 |
| Social Function - YR2 | Overall Health - BL | 0.00 | 0.00 | 0.01 | 0.00 | 0.00 | 0.00 | 0.99 |
| Overall Health - YR3 | Physical Limit - YR4 | 0.00 | 0.01 | 0.00 | 0.00 | 0.00 | 0.00 | 0.99 |
| Depression - YR3 | Quality of Life - YR4 | 0.00 | 0.01 | 0.00 | 0.00 | 0.00 | 0.00 | 0.99 |
| PTSD Symptoms - YR5 | Depression - BL | 0.00 | 0.00 | 0.01 | 0.00 | 0.00 | 0.00 | 0.99 |
| Physical Limit - YR5 | Depression - YR5 | 0.01 | 0.00 | 0.00 | 0.00 | 0.00 | 0.00 | 0.99 |
| Overall Health - YR6 | Overall Health - BL | 0.00 | 0.00 | 0.01 | 0.00 | 0.00 | 0.00 | 0.99 |
| Depression - YR7 | Quality of Life - YR7 | 0.01 | 0.00 | 0.00 | 0.00 | 0.00 | 0.00 | 0.99 |
| Social Function - YR7 | Quality of Life - BL | 0.00 | 0.00 | 0.01 | 0.00 | 0.00 | 0.00 | 0.99 |
| Overall Health - YR4 | Depression - YR4 | 0.01 | 0.00 | 0.00 | 0.00 | 0.00 | 0.00 | 0.99 |
| Overall Health - YR1 | Physical Amount - YR6 | 0.00 | 0.01 | 0.00 | 0.00 | 0.00 | 0.00 | 0.99 |
| Physical Compared - YR1 | Physical Compared - YR5 | 0.00 | 0.01 | 0.00 | 0.00 | 0.00 | 0.00 | 0.99 |
| Physical Amount - YR2 | Social Function - BL | 0.00 | 0.00 | 0.01 | 0.00 | 0.00 | 0.00 | 0.99 |
| Physical Compared - YR2 | Physical Limit - YR2 | 0.01 | 0.00 | 0.00 | 0.00 | 0.00 | 0.00 | 0.99 |
| Overall Health - YR5 | Depression - YR5 | 0.00 | 0.01 | 0.00 | 0.00 | 0.00 | 0.00 | 0.99 |
| Depression - YR6 | Depression - BL | 0.00 | 0.00 | 0.01 | 0.00 | 0.00 | 0.00 | 0.99 |
| Physical Amount - YR7 | Physical Limit - BL | 0.00 | 0.00 | 0.01 | 0.00 | 0.00 | 0.00 | 0.99 |
| Quality of Life - BL | Quality of Life - YR4 | 0.00 | 0.00 | 0.00 | 0.01 | 0.00 | 0.00 | 0.99 |
| Overall Health - YR1 | Social Function - YR1 | 0.00 | 0.00 | 0.01 | 0.00 | 0.00 | 0.00 | 0.99 |
| PTSD Symptoms - YR1 | Physical Amount - YR7 | 0.00 | 0.01 | 0.00 | 0.00 | 0.00 | 0.00 | 0.99 |
| Depression - YR1 | PTSD Symptoms - YR5 | 0.00 | 0.01 | 0.00 | 0.00 | 0.00 | 0.00 | 0.99 |
| Depression - YR1 | Depression - YR6 | 0.00 | 0.01 | 0.00 | 0.00 | 0.00 | 0.00 | 0.99 |
| Social Function - YR1 | Physical Amount - YR4 | 0.00 | 0.00 | 0.00 | 0.00 | 0.00 | 0.00 | 0.99 |
| PTSD Symptoms - YR2 | PTSD Symptoms - YR5 | 0.00 | 0.01 | 0.00 | 0.00 | 0.00 | 0.00 | 0.99 |
| PTSD Symptoms - YR2 | Physical Limit - YR2 | 0.00 | 0.01 | 0.00 | 0.00 | 0.00 | 0.00 | 0.99 |
| Physical Amount - YR3 | Depression - BL | 0.00 | 0.00 | 0.01 | 0.00 | 0.00 | 0.00 | 0.99 |
| Social Function - YR3 | Overall Health - BL | 0.00 | 0.00 | 0.01 | 0.00 | 0.00 | 0.00 | 0.99 |
| PTSD Symptoms - YR6 | Depression - BL | 0.00 | 0.00 | 0.01 | 0.00 | 0.00 | 0.00 | 0.99 |
| PTSD Symptoms - YR7 | Depression - BL | 0.00 | 0.00 | 0.01 | 0.00 | 0.00 | 0.00 | 0.99 |
| Physical Limit - BL | Physical Limit - YR4 | 0.00 | 0.00 | 0.00 | 0.01 | 0.00 | 0.00 | 0.99 |
| Alcohol Use - BL | Quality of Life - BL | 0.00 | 0.00 | 0.00 | 0.00 | 0.01 | 0.00 | 1.00 |
| Overall Health - YR4 | Depression - YR5 | 0.00 | 0.01 | 0.00 | 0.00 | 0.00 | 0.00 | 1.00 |

Table S3. Stability analysis results on the MYH data

| Variable 1 | Variable 2 | <-- | --> | <-o | o-> | o-o | <-> | No Edge |
|---|---|---|---|---|---|---|---|---|
| Physical Amount - YR2 | Physical Limit - BL | 0.00 | 0.00 | 0.01 | 0.00 | 0.00 | 0.00 | 1.00 |
| Depression - YR2 | PTSD Symptoms - YR3 | 0.00 | 0.01 | 0.00 | 0.00 | 0.00 | 0.00 | 1.00 |
| Quality of Life - YR2 | Quality of Life - YR6 | 0.00 | 0.01 | 0.00 | 0.00 | 0.00 | 0.00 | 1.00 |
| Quality of Life - YR2 | Depression - YR6 | 0.00 | 0.01 | 0.00 | 0.00 | 0.00 | 0.00 | 1.00 |
| PTSD Symptoms - YR3 | Social Function - BL | 0.00 | 0.00 | 0.01 | 0.00 | 0.00 | 0.00 | 1.00 |
| Physical Amount - YR3 | Social Function - BL | 0.00 | 0.00 | 0.01 | 0.00 | 0.00 | 0.00 | 1.00 |
| Social Function - YR3 | PTSD Symptoms - YR5 | 0.00 | 0.01 | 0.00 | 0.00 | 0.00 | 0.00 | 1.00 |
| Depression - YR4 | Depression - BL | 0.00 | 0.00 | 0.01 | 0.00 | 0.00 | 0.00 | 1.00 |
| Physical Compared - YR6 | Overall Health - BL | 0.00 | 0.00 | 0.01 | 0.00 | 0.00 | 0.00 | 1.00 |
| Quality of Life - YR6 | Overall Health - YR7 | 0.00 | 0.01 | 0.00 | 0.00 | 0.00 | 0.00 | 1.00 |
| Overall Health - YR4 | Social Function - YR3 | 0.00 | 0.00 | 0.00 | 0.00 | 0.00 | 0.00 | 1.00 |
| Overall Health - YR4 | Physical Limit - YR3 | 0.00 | 0.00 | 0.00 | 0.00 | 0.00 | 0.00 | 1.00 |
| Overall Health - YR4 | Physical Compared - YR7 | 0.00 | 0.00 | 0.00 | 0.00 | 0.00 | 0.00 | 1.00 |
| Overall Health - YR1 | Physical Amount - YR1 | 0.00 | 0.00 | 0.00 | 0.00 | 0.00 | 0.00 | 1.00 |
| Physical Amount - YR1 | Social Function - YR1 | 0.00 | 0.00 | 0.00 | 0.00 | 0.00 | 0.00 | 1.00 |
| Depression - YR1 | Physical Amount - YR2 | 0.00 | 0.00 | 0.00 | 0.00 | 0.00 | 0.00 | 1.00 |
| Depression - YR1 | Social Function - YR1 | 0.00 | 0.00 | 0.00 | 0.00 | 0.00 | 0.00 | 1.00 |
| Quality of Life - YR1 | Physical Amount - YR5 | 0.00 | 0.00 | 0.00 | 0.00 | 0.00 | 0.00 | 1.00 |
| Overall Health - YR2 | Physical Limit - YR7 | 0.00 | 0.00 | 0.00 | 0.00 | 0.00 | 0.00 | 1.00 |
| Depression - YR2 | Depression - YR5 | 0.00 | 0.00 | 0.00 | 0.00 | 0.00 | 0.00 | 1.00 |
| Depression - YR2 | PTSD Symptoms - YR5 | 0.00 | 0.00 | 0.00 | 0.00 | 0.00 | 0.00 | 1.00 |
| Quality of Life - YR2 | Depression - YR4 | 0.00 | 0.00 | 0.00 | 0.00 | 0.00 | 0.00 | 1.00 |
| Depression - YR3 | PTSD Symptoms - YR5 | 0.00 | 0.00 | 0.00 | 0.00 | 0.00 | 0.00 | 1.00 |
| Quality of Life - YR3 | Overall Health - YR6 | 0.00 | 0.00 | 0.00 | 0.00 | 0.00 | 0.00 | 1.00 |
| Social Function - YR3 | Quality of Life - BL | 0.00 | 0.00 | 0.00 | 0.00 | 0.00 | 0.00 | 1.00 |
| Depression - YR4 | Depression - YR6 | 0.00 | 0.00 | 0.00 | 0.00 | 0.00 | 0.00 | 1.00 |
| Physical Limit - YR5 | Quality of Life - YR5 | 0.00 | 0.00 | 0.00 | 0.00 | 0.00 | 0.00 | 1.00 |
| Social Function - YR5 | Depression - YR6 | 0.00 | 0.00 | 0.00 | 0.00 | 0.00 | 0.00 | 1.00 |
| Social Function - YR5 | Social Function - BL | 0.00 | 0.00 | 0.00 | 0.00 | 0.00 | 0.00 | 1.00 |
| Physical Amount - YR6 | Overall Health - BL | 0.00 | 0.00 | 0.00 | 0.00 | 0.00 | 0.00 | 1.00 |
| Physical Compared - YR6 | Physical Limit - YR6 | 0.00 | 0.00 | 0.00 | 0.00 | 0.00 | 0.00 | 1.00 |
| Physical Limit - YR6 | Physical Limit - BL | 0.00 | 0.00 | 0.00 | 0.00 | 0.00 | 0.00 | 1.00 |
| Physical Amount - YR7 | Social Function - BL | 0.00 | 0.00 | 0.00 | 0.00 | 0.00 | 0.00 | 1.00 |
| Physical Compared - YR7 | Depression - YR7 | 0.00 | 0.00 | 0.00 | 0.00 | 0.00 | 0.00 | 1.00 |
| Overall Health - YR1 | Physical Compared - YR5 | 0.00 | 0.00 | 0.00 | 0.00 | 0.00 | 0.00 | 1.00 |
| PTSD Symptoms - YR1 | Quality of Life - YR1 | 0.00 | 0.00 | 0.00 | 0.00 | 0.00 | 0.00 | 1.00 |
| Physical Compared - YR1 | Quality of Life - BL | 0.00 | 0.00 | 0.00 | 0.00 | 0.00 | 0.00 | 1.00 |
| Physical Limit - YR1 | Overall Health - YR2 | 0.00 | 0.00 | 0.00 | 0.00 | 0.00 | 0.00 | 1.00 |
| Depression - YR1 | Depression - YR3 | 0.00 | 0.00 | 0.00 | 0.00 | 0.00 | 0.00 | 1.00 |
| Depression - YR1 | Social Function - BL | 0.00 | 0.00 | 0.00 | 0.00 | 0.00 | 0.00 | 1.00 |
| Social Function - YR1 | Depression - YR4 | 0.00 | 0.00 | 0.00 | 0.00 | 0.00 | 0.00 | 1.00 |
| Overall Health - YR2 | Overall Health - YR7 | 0.00 | 0.00 | 0.00 | 0.00 | 0.00 | 0.00 | 1.00 |
| Depression - YR2 | Physical Compared - YR3 | 0.00 | 0.00 | 0.00 | 0.00 | 0.00 | 0.00 | 1.00 |
| PTSD Symptoms - YR3 | Physical Amount - YR3 | 0.00 | 0.00 | 0.00 | 0.00 | 0.00 | 0.00 | 1.00 |
| PTSD Symptoms - YR3 | Physical Limit - YR4 | 0.00 | 0.00 | 0.00 | 0.00 | 0.00 | 0.00 | 1.00 |
| Physical Compared - YR3 | Overall Health - YR6 | 0.00 | 0.00 | 0.00 | 0.00 | 0.00 | 0.00 | 1.00 |
| Physical Limit - YR3 | Depression - YR4 | 0.00 | 0.00 | 0.00 | 0.00 | 0.00 | 0.00 | 1.00 |

Table S3. Stability analysis results on the MYH data

| Variable 1 | Variable 2 | <-- | --> | <-o | o-> | o-o | <-> | No Edge |
|---|---|---|---|---|---|---|---|---|
| Depression - YR3 | Physical Limit - YR7 | 0.00 | 0.00 | 0.00 | 0.00 | 0.00 | 0.00 | 1.00 |
| Overall Health - YR5 | Physical Compared - YR7 | 0.00 | 0.00 | 0.00 | 0.00 | 0.00 | 0.00 | 1.00 |
| PTSD Symptoms - YR5 | Physical Limit - YR6 | 0.00 | 0.00 | 0.00 | 0.00 | 0.00 | 0.00 | 1.00 |
| Physical Amount - YR5 | Social Function - BL | 0.00 | 0.00 | 0.00 | 0.00 | 0.00 | 0.00 | 1.00 |
| Depression - YR5 | Overall Health - YR6 | 0.00 | 0.00 | 0.00 | 0.00 | 0.00 | 0.00 | 1.00 |
| Quality of Life - YR5 | Quality of Life - BL | 0.00 | 0.00 | 0.00 | 0.00 | 0.00 | 0.00 | 1.00 |
| Social Function - YR5 | Overall Health - YR7 | 0.00 | 0.00 | 0.00 | 0.00 | 0.00 | 0.00 | 1.00 |
| Social Function - YR5 | Physical Limit - YR6 | 0.00 | 0.00 | 0.00 | 0.00 | 0.00 | 0.00 | 1.00 |
| Physical Amount - YR6 | PTSD Symptoms - BL | 0.00 | 0.00 | 0.00 | 0.00 | 0.00 | 0.00 | 1.00 |
| Physical Compared - YR6 | Quality of Life - BL | 0.00 | 0.00 | 0.00 | 0.00 | 0.00 | 0.00 | 1.00 |
| Physical Compared - YR7 | Physical Limit - YR4 | 0.00 | 0.00 | 0.00 | 0.00 | 0.00 | 0.00 | 1.00 |
| Alcohol Use - YR1 | Depression - YR1 | 0.00 | 0.00 | 0.00 | 0.00 | 0.00 | 0.00 | 1.00 |
| Overall Health - YR1 | PTSD Symptoms - YR3 | 0.00 | 0.00 | 0.00 | 0.00 | 0.00 | 0.00 | 1.00 |
| Overall Health - YR1 | Quality of Life - YR2 | 0.00 | 0.00 | 0.00 | 0.00 | 0.00 | 0.00 | 1.00 |
| Overall Health - YR1 | Overall Health - YR6 | 0.00 | 0.00 | 0.00 | 0.00 | 0.00 | 0.00 | 1.00 |
| Overall Health - YR1 | Physical Amount - YR7 | 0.00 | 0.00 | 0.00 | 0.00 | 0.00 | 0.00 | 1.00 |
| PTSD Symptoms - YR1 | Overall Health - YR3 | 0.00 | 0.00 | 0.00 | 0.00 | 0.00 | 0.00 | 1.00 |
| PTSD Symptoms - YR1 | Physical Amount - YR5 | 0.00 | 0.00 | 0.00 | 0.00 | 0.00 | 0.00 | 1.00 |
| PTSD Symptoms - YR1 | Physical Limit - BL | 0.00 | 0.00 | 0.00 | 0.00 | 0.00 | 0.00 | 1.00 |
| Physical Amount - YR1 | Physical Compared - YR2 | 0.00 | 0.00 | 0.00 | 0.00 | 0.00 | 0.00 | 1.00 |
| Physical Amount - YR1 | Overall Health - BL | 0.00 | 0.00 | 0.00 | 0.00 | 0.00 | 0.00 | 1.00 |
| Physical Limit - YR1 | Physical Compared - YR7 | 0.00 | 0.00 | 0.00 | 0.00 | 0.00 | 0.00 | 1.00 |
| Physical Limit - YR1 | Physical Compared - YR4 | 0.00 | 0.00 | 0.00 | 0.00 | 0.00 | 0.00 | 1.00 |
| Depression - YR1 | Depression - YR2 | 0.00 | 0.00 | 0.00 | 0.00 | 0.00 | 0.00 | 1.00 |
| Depression - YR1 | PTSD Symptoms - YR2 | 0.00 | 0.00 | 0.00 | 0.00 | 0.00 | 0.00 | 1.00 |
| Depression - YR1 | Physical Amount - YR6 | 0.00 | 0.00 | 0.00 | 0.00 | 0.00 | 0.00 | 1.00 |
| Social Function - YR1 | Physical Amount - YR2 | 0.00 | 0.00 | 0.00 | 0.00 | 0.00 | 0.00 | 1.00 |
| Alcohol Use - YR2 | Physical Limit - BL | 0.00 | 0.00 | 0.00 | 0.00 | 0.00 | 0.00 | 1.00 |
| Overall Health - YR2 | PTSD Symptoms - YR3 | 0.00 | 0.00 | 0.00 | 0.00 | 0.00 | 0.00 | 1.00 |
| Overall Health - YR2 | Physical Limit - YR5 | 0.00 | 0.00 | 0.00 | 0.00 | 0.00 | 0.00 | 1.00 |
| Quality of Life - YR2 | PTSD Symptoms - YR4 | 0.00 | 0.00 | 0.00 | 0.00 | 0.00 | 0.00 | 1.00 |
| Quality of Life - YR2 | Physical Limit - YR3 | 0.00 | 0.00 | 0.00 | 0.00 | 0.00 | 0.00 | 1.00 |
| Quality of Life - YR2 | Depression - YR7 | 0.00 | 0.00 | 0.00 | 0.00 | 0.00 | 0.00 | 1.00 |
| Social Function - YR2 | PTSD Symptoms - YR6 | 0.00 | 0.00 | 0.00 | 0.00 | 0.00 | 0.00 | 1.00 |
| Social Function - YR2 | Physical Limit - YR3 | 0.00 | 0.00 | 0.00 | 0.00 | 0.00 | 0.00 | 1.00 |
| Social Function - YR2 | Quality of Life - YR6 | 0.00 | 0.00 | 0.00 | 0.00 | 0.00 | 0.00 | 1.00 |
| Overall Health - YR3 | Physical Compared - YR5 | 0.00 | 0.00 | 0.00 | 0.00 | 0.00 | 0.00 | 1.00 |
| Overall Health - YR3 | Physical Limit - YR5 | 0.00 | 0.00 | 0.00 | 0.00 | 0.00 | 0.00 | 1.00 |
| Physical Compared - YR3 | Quality of Life - YR3 | 0.00 | 0.00 | 0.00 | 0.00 | 0.00 | 0.00 | 1.00 |
| Physical Compared - YR3 | Depression - YR6 | 0.00 | 0.00 | 0.00 | 0.00 | 0.00 | 0.00 | 1.00 |
| Physical Limit - YR3 | Social Function - BL | 0.00 | 0.00 | 0.00 | 0.00 | 0.00 | 0.00 | 1.00 |
| Quality of Life - YR3 | Overall Health - YR7 | 0.00 | 0.00 | 0.00 | 0.00 | 0.00 | 0.00 | 1.00 |
| Depression - YR4 | Depression - YR7 | 0.00 | 0.00 | 0.00 | 0.00 | 0.00 | 0.00 | 1.00 |
| PTSD Symptoms - YR5 | Quality of Life - YR5 | 0.00 | 0.00 | 0.00 | 0.00 | 0.00 | 0.00 | 1.00 |
| PTSD Symptoms - YR5 | Physical Compared - YR5 | 0.00 | 0.00 | 0.00 | 0.00 | 0.00 | 0.00 | 1.00 |
| Physical Amount - YR5 | Physical Limit - YR6 | 0.00 | 0.00 | 0.00 | 0.00 | 0.00 | 0.00 | 1.00 |
| Physical Limit - YR5 | Overall Health - YR6 | 0.00 | 0.00 | 0.00 | 0.00 | 0.00 | 0.00 | 1.00 |

Table S3. Stability analysis results on the MYH data

| Variable 1 | Variable 2 | <-- | --> | <-o | o-> | o-o | <-> | No Edge |
|---|---|---|---|---|---|---|---|---|
| Social Function - YR5 | Overall Health - YR6 | 0.00 | 0.00 | 0.00 | 0.00 | 0.00 | 0.00 | 1.00 |
| PTSD Symptoms - YR6 | Social Function - YR4 | 0.00 | 0.00 | 0.00 | 0.00 | 0.00 | 0.00 | 1.00 |
| Physical Amount - YR6 | Quality of Life - BL | 0.00 | 0.00 | 0.00 | 0.00 | 0.00 | 0.00 | 1.00 |
| Physical Amount - YR6 | Depression - BL | 0.00 | 0.00 | 0.00 | 0.00 | 0.00 | 0.00 | 1.00 |
| Physical Compared - YR6 | Physical Limit - YR4 | 0.00 | 0.00 | 0.00 | 0.00 | 0.00 | 0.00 | 1.00 |
| Physical Limit - YR7 | Depression - BL | 0.00 | 0.00 | 0.00 | 0.00 | 0.00 | 0.00 | 1.00 |
| PTSD Symptoms - BL | Physical Amount - YR4 | 0.00 | 0.00 | 0.00 | 0.00 | 0.00 | 0.00 | 1.00 |
| PTSD Symptoms - BL | Physical Limit - YR4 | 0.00 | 0.00 | 0.00 | 0.00 | 0.00 | 0.00 | 1.00 |
| Quality of Life - YR4 | Social Function - YR4 | 0.00 | 0.00 | 0.00 | 0.00 | 0.00 | 0.00 | 1.00 |
| Alcohol Use - BL | Social Function - YR2 | 0.00 | 0.00 | 0.00 | 0.00 | 0.00 | 0.00 | 1.00 |
| Alcohol Use - BL | Physical Amount - YR6 | 0.00 | 0.00 | 0.00 | 0.00 | 0.00 | 0.00 | 1.00 |
| Alcohol Use - YR4 | PTSD Symptoms - YR7 | 0.00 | 0.00 | 0.00 | 0.00 | 0.00 | 0.00 | 1.00 |
| Alcohol Use - YR4 | Social Function - BL | 0.00 | 0.00 | 0.00 | 0.00 | 0.00 | 0.00 | 1.00 |
| Overall Health - YR4 | Physical Compared - YR6 | 0.00 | 0.00 | 0.00 | 0.00 | 0.00 | 0.00 | 1.00 |
| Overall Health - YR4 | Quality of Life - YR2 | 0.00 | 0.00 | 0.00 | 0.00 | 0.00 | 0.00 | 1.00 |
| Overall Health - YR4 | Social Function - YR2 | 0.00 | 0.00 | 0.00 | 0.00 | 0.00 | 0.00 | 1.00 |
| Overall Health - YR4 | Physical Compared - YR5 | 0.00 | 0.00 | 0.00 | 0.00 | 0.00 | 0.00 | 1.00 |
| Overall Health - YR4 | Physical Limit - YR6 | 0.00 | 0.00 | 0.00 | 0.00 | 0.00 | 0.00 | 1.00 |
| Overall Health - YR4 | Quality of Life - YR7 | 0.00 | 0.00 | 0.00 | 0.00 | 0.00 | 0.00 | 1.00 |
| Overall Health - YR4 | Physical Limit - BL | 0.00 | 0.00 | 0.00 | 0.00 | 0.00 | 0.00 | 1.00 |
| Alcohol Use - YR1 | Physical Amount - YR2 | 0.00 | 0.00 | 0.00 | 0.00 | 0.00 | 0.00 | 1.00 |
| Alcohol Use - YR1 | Physical Limit - BL | 0.00 | 0.00 | 0.00 | 0.00 | 0.00 | 0.00 | 1.00 |
| Overall Health - YR1 | Quality of Life - YR3 | 0.00 | 0.00 | 0.00 | 0.00 | 0.00 | 0.00 | 1.00 |
| Overall Health - YR1 | Physical Limit - YR5 | 0.00 | 0.00 | 0.00 | 0.00 | 0.00 | 0.00 | 1.00 |
| Overall Health - YR1 | Physical Amount - YR4 | 0.00 | 0.00 | 0.00 | 0.00 | 0.00 | 0.00 | 1.00 |
| Overall Health - YR1 | Physical Limit - YR4 | 0.00 | 0.00 | 0.00 | 0.00 | 0.00 | 0.00 | 1.00 |
| PTSD Symptoms - YR1 | Physical Amount - YR3 | 0.00 | 0.00 | 0.00 | 0.00 | 0.00 | 0.00 | 1.00 |
| PTSD Symptoms - YR1 | Physical Amount - YR2 | 0.00 | 0.00 | 0.00 | 0.00 | 0.00 | 0.00 | 1.00 |
| PTSD Symptoms - YR1 | Physical Amount - YR1 | 0.00 | 0.00 | 0.00 | 0.00 | 0.00 | 0.00 | 1.00 |
| Physical Amount - YR1 | PTSD Symptoms - YR3 | 0.00 | 0.00 | 0.00 | 0.00 | 0.00 | 0.00 | 1.00 |
| Physical Amount - YR1 | Depression - YR2 | 0.00 | 0.00 | 0.00 | 0.00 | 0.00 | 0.00 | 1.00 |
| Physical Amount - YR1 | Physical Compared - YR3 | 0.00 | 0.00 | 0.00 | 0.00 | 0.00 | 0.00 | 1.00 |
| Physical Amount - YR1 | PTSD Symptoms - YR4 | 0.00 | 0.00 | 0.00 | 0.00 | 0.00 | 0.00 | 1.00 |
| Physical Amount - YR1 | Physical Compared - YR4 | 0.00 | 0.00 | 0.00 | 0.00 | 0.00 | 0.00 | 1.00 |
| Physical Limit - YR1 | Physical Compared - YR3 | 0.00 | 0.00 | 0.00 | 0.00 | 0.00 | 0.00 | 1.00 |
| Physical Limit - YR1 | Overall Health - YR7 | 0.00 | 0.00 | 0.00 | 0.00 | 0.00 | 0.00 | 1.00 |
| Depression - YR1 | Depression - YR4 | 0.00 | 0.00 | 0.00 | 0.00 | 0.00 | 0.00 | 1.00 |
| Depression - YR1 | Physical Amount - YR3 | 0.00 | 0.00 | 0.00 | 0.00 | 0.00 | 0.00 | 1.00 |
| Depression - YR1 | Overall Health - YR2 | 0.00 | 0.00 | 0.00 | 0.00 | 0.00 | 0.00 | 1.00 |
| Depression - YR1 | Depression - YR5 | 0.00 | 0.00 | 0.00 | 0.00 | 0.00 | 0.00 | 1.00 |
| Depression - YR1 | Quality of Life - YR6 | 0.00 | 0.00 | 0.00 | 0.00 | 0.00 | 0.00 | 1.00 |
| Depression - YR1 | Quality of Life - BL | 0.00 | 0.00 | 0.00 | 0.00 | 0.00 | 0.00 | 1.00 |
| Quality of Life - YR1 | PTSD Symptoms - YR6 | 0.00 | 0.00 | 0.00 | 0.00 | 0.00 | 0.00 | 1.00 |
| Quality of Life - YR1 | Physical Amount - YR2 | 0.00 | 0.00 | 0.00 | 0.00 | 0.00 | 0.00 | 1.00 |
| Quality of Life - YR1 | Physical Amount - YR3 | 0.00 | 0.00 | 0.00 | 0.00 | 0.00 | 0.00 | 1.00 |
| Quality of Life - YR1 | Depression - YR6 | 0.00 | 0.00 | 0.00 | 0.00 | 0.00 | 0.00 | 1.00 |
| Quality of Life - YR1 | Physical Compared - YR6 | 0.00 | 0.00 | 0.00 | 0.00 | 0.00 | 0.00 | 1.00 |

Table S3. Stability analysis results on the MYH data

| Variable 1 | Variable 2 | <-- | --> | <-o | o-> | o-o | <-> | No Edge |
|---|---|---|---|---|---|---|---|---|
| Quality of Life - YR1 | Depression - BL | 0.00 | 0.00 | 0.00 | 0.00 | 0.00 | 0.00 | 1.00 |
| Social Function - YR1 | Overall Health - YR3 | 0.00 | 0.00 | 0.00 | 0.00 | 0.00 | 0.00 | 1.00 |
| Social Function - YR1 | Depression - YR2 | 0.00 | 0.00 | 0.00 | 0.00 | 0.00 | 0.00 | 1.00 |
| Alcohol Use - YR2 | Social Function - BL | 0.00 | 0.00 | 0.00 | 0.00 | 0.00 | 0.00 | 1.00 |
| Overall Health - YR2 | Physical Amount - YR2 | 0.00 | 0.00 | 0.00 | 0.00 | 0.00 | 0.00 | 1.00 |
| Overall Health - YR2 | Quality of Life - YR5 | 0.00 | 0.00 | 0.00 | 0.00 | 0.00 | 0.00 | 1.00 |
| Overall Health - YR2 | Physical Compared - YR6 | 0.00 | 0.00 | 0.00 | 0.00 | 0.00 | 0.00 | 1.00 |
| Overall Health - YR2 | PTSD Symptoms - BL | 0.00 | 0.00 | 0.00 | 0.00 | 0.00 | 0.00 | 1.00 |
| PTSD Symptoms - YR2 | Quality of Life - YR2 | 0.00 | 0.00 | 0.00 | 0.00 | 0.00 | 0.00 | 1.00 |
| PTSD Symptoms - YR2 | Depression - YR7 | 0.00 | 0.00 | 0.00 | 0.00 | 0.00 | 0.00 | 1.00 |
| PTSD Symptoms - YR2 | Depression - BL | 0.00 | 0.00 | 0.00 | 0.00 | 0.00 | 0.00 | 1.00 |
| Physical Amount - YR2 | Overall Health - YR6 | 0.00 | 0.00 | 0.00 | 0.00 | 0.00 | 0.00 | 1.00 |
| Depression - YR2 | Physical Limit - YR3 | 0.00 | 0.00 | 0.00 | 0.00 | 0.00 | 0.00 | 1.00 |
| Depression - YR2 | Social Function - BL | 0.00 | 0.00 | 0.00 | 0.00 | 0.00 | 0.00 | 1.00 |
| Quality of Life - YR2 | Physical Compared - YR3 | 0.00 | 0.00 | 0.00 | 0.00 | 0.00 | 0.00 | 1.00 |
| Quality of Life - YR2 | Overall Health - YR7 | 0.00 | 0.00 | 0.00 | 0.00 | 0.00 | 0.00 | 1.00 |
| Social Function - YR2 | Physical Compared - YR3 | 0.00 | 0.00 | 0.00 | 0.00 | 0.00 | 0.00 | 1.00 |
| Social Function - YR2 | Overall Health - YR3 | 0.00 | 0.00 | 0.00 | 0.00 | 0.00 | 0.00 | 1.00 |
| Social Function - YR2 | Quality of Life - BL | 0.00 | 0.00 | 0.00 | 0.00 | 0.00 | 0.00 | 1.00 |
| PTSD Symptoms - YR3 | Physical Limit - YR3 | 0.00 | 0.00 | 0.00 | 0.00 | 0.00 | 0.00 | 1.00 |
| Physical Compared - YR3 | Overall Health - YR5 | 0.00 | 0.00 | 0.00 | 0.00 | 0.00 | 0.00 | 1.00 |
| Physical Compared - YR3 | Physical Limit - YR5 | 0.00 | 0.00 | 0.00 | 0.00 | 0.00 | 0.00 | 1.00 |
| Physical Limit - YR3 | Overall Health - YR6 | 0.00 | 0.00 | 0.00 | 0.00 | 0.00 | 0.00 | 1.00 |
| Physical Limit - YR3 | Physical Compared - YR7 | 0.00 | 0.00 | 0.00 | 0.00 | 0.00 | 0.00 | 1.00 |
| Physical Limit - YR3 | Overall Health - BL | 0.00 | 0.00 | 0.00 | 0.00 | 0.00 | 0.00 | 1.00 |
| Quality of Life - YR3 | Depression - YR4 | 0.00 | 0.00 | 0.00 | 0.00 | 0.00 | 0.00 | 1.00 |
| Quality of Life - YR3 | Physical Compared - YR6 | 0.00 | 0.00 | 0.00 | 0.00 | 0.00 | 0.00 | 1.00 |
| Quality of Life - YR3 | Physical Compared - YR4 | 0.00 | 0.00 | 0.00 | 0.00 | 0.00 | 0.00 | 1.00 |
| Quality of Life - YR3 | Physical Limit - YR4 | 0.00 | 0.00 | 0.00 | 0.00 | 0.00 | 0.00 | 1.00 |
| Social Function - YR3 | Overall Health - YR6 | 0.00 | 0.00 | 0.00 | 0.00 | 0.00 | 0.00 | 1.00 |
| PTSD Symptoms - YR4 | Overall Health - YR7 | 0.00 | 0.00 | 0.00 | 0.00 | 0.00 | 0.00 | 1.00 |
| PTSD Symptoms - YR4 | Social Function - BL | 0.00 | 0.00 | 0.00 | 0.00 | 0.00 | 0.00 | 1.00 |
| Depression - YR4 | PTSD Symptoms - YR5 | 0.00 | 0.00 | 0.00 | 0.00 | 0.00 | 0.00 | 1.00 |
| Alcohol Use - YR5 | Social Function - BL | 0.00 | 0.00 | 0.00 | 0.00 | 0.00 | 0.00 | 1.00 |
| Overall Health - YR5 | Physical Limit - YR5 | 0.00 | 0.00 | 0.00 | 0.00 | 0.00 | 0.00 | 1.00 |
| Overall Health - YR5 | Physical Limit - YR7 | 0.00 | 0.00 | 0.00 | 0.00 | 0.00 | 0.00 | 1.00 |
| PTSD Symptoms - YR5 | Overall Health - YR6 | 0.00 | 0.00 | 0.00 | 0.00 | 0.00 | 0.00 | 1.00 |
| PTSD Symptoms - YR5 | Quality of Life - YR7 | 0.00 | 0.00 | 0.00 | 0.00 | 0.00 | 0.00 | 1.00 |
| Physical Compared - YR5 | Physical Limit - YR6 | 0.00 | 0.00 | 0.00 | 0.00 | 0.00 | 0.00 | 1.00 |
| Physical Compared - YR5 | Physical Amount - YR4 | 0.00 | 0.00 | 0.00 | 0.00 | 0.00 | 0.00 | 1.00 |
| Physical Limit - YR5 | PTSD Symptoms - YR7 | 0.00 | 0.00 | 0.00 | 0.00 | 0.00 | 0.00 | 1.00 |
| Physical Limit - YR5 | Social Function - BL | 0.00 | 0.00 | 0.00 | 0.00 | 0.00 | 0.00 | 1.00 |
| Depression - YR5 | Physical Limit - YR7 | 0.00 | 0.00 | 0.00 | 0.00 | 0.00 | 0.00 | 1.00 |
| Depression - YR5 | PTSD Symptoms - YR7 | 0.00 | 0.00 | 0.00 | 0.00 | 0.00 | 0.00 | 1.00 |
| Depression - YR5 | PTSD Symptoms - YR6 | 0.00 | 0.00 | 0.00 | 0.00 | 0.00 | 0.00 | 1.00 |
| Depression - YR5 | Overall Health - BL | 0.00 | 0.00 | 0.00 | 0.00 | 0.00 | 0.00 | 1.00 |
| Depression - YR5 | Social Function - BL | 0.00 | 0.00 | 0.00 | 0.00 | 0.00 | 0.00 | 1.00 |

Table S3. Stability analysis results on the MYH data

| Variable 1 | Variable 2 | <-- | --> | <-o | o-> | o-o | <-> | No Edge |
|---|---|---|---|---|---|---|---|---|
| Depression - YR5 | Social Function - YR4 | 0.00 | 0.00 | 0.00 | 0.00 | 0.00 | 0.00 | 1.00 |
| Depression - YR5 | Quality of Life - YR4 | 0.00 | 0.00 | 0.00 | 0.00 | 0.00 | 0.00 | 1.00 |
| Quality of Life - YR5 | Depression - YR6 | 0.00 | 0.00 | 0.00 | 0.00 | 0.00 | 0.00 | 1.00 |
| Social Function - YR5 | PTSD Symptoms - YR7 | 0.00 | 0.00 | 0.00 | 0.00 | 0.00 | 0.00 | 1.00 |
| Overall Health - YR6 | Physical Limit - YR7 | 0.00 | 0.00 | 0.00 | 0.00 | 0.00 | 0.00 | 1.00 |
| Overall Health - YR6 | Physical Limit - BL | 0.00 | 0.00 | 0.00 | 0.00 | 0.00 | 0.00 | 1.00 |
| Overall Health - YR6 | Social Function - BL | 0.00 | 0.00 | 0.00 | 0.00 | 0.00 | 0.00 | 1.00 |
| Overall Health - YR6 | Social Function - YR4 | 0.00 | 0.00 | 0.00 | 0.00 | 0.00 | 0.00 | 1.00 |
| PTSD Symptoms - YR6 | Physical Limit - BL | 0.00 | 0.00 | 0.00 | 0.00 | 0.00 | 0.00 | 1.00 |
| Physical Amount - YR6 | Social Function - BL | 0.00 | 0.00 | 0.00 | 0.00 | 0.00 | 0.00 | 1.00 |
| Physical Compared - YR6 | Quality of Life - YR4 | 0.00 | 0.00 | 0.00 | 0.00 | 0.00 | 0.00 | 1.00 |
| Quality of Life - YR6 | Depression - YR7 | 0.00 | 0.00 | 0.00 | 0.00 | 0.00 | 0.00 | 1.00 |
| Quality of Life - YR6 | Quality of Life - BL | 0.00 | 0.00 | 0.00 | 0.00 | 0.00 | 0.00 | 1.00 |
| Social Function - YR6 | Physical Limit - BL | 0.00 | 0.00 | 0.00 | 0.00 | 0.00 | 0.00 | 1.00 |
| Alcohol Use - YR7 | PTSD Symptoms - YR7 | 0.00 | 0.00 | 0.00 | 0.00 | 0.00 | 0.00 | 1.00 |
| Overall Health - YR7 | Quality of Life - YR4 | 0.00 | 0.00 | 0.00 | 0.00 | 0.00 | 0.00 | 1.00 |
| PTSD Symptoms - YR7 | Physical Limit - YR4 | 0.00 | 0.00 | 0.00 | 0.00 | 0.00 | 0.00 | 1.00 |
| Physical Amount - YR7 | Physical Limit - YR7 | 0.00 | 0.00 | 0.00 | 0.00 | 0.00 | 0.00 | 1.00 |
| Physical Limit - YR7 | Physical Limit - YR4 | 0.00 | 0.00 | 0.00 | 0.00 | 0.00 | 0.00 | 1.00 |
| Quality of Life - YR7 | Depression - BL | 0.00 | 0.00 | 0.00 | 0.00 | 0.00 | 0.00 | 1.00 |
| Social Function - YR7 | Physical Limit - BL | 0.00 | 0.00 | 0.00 | 0.00 | 0.00 | 0.00 | 1.00 |
| Social Function - YR7 | Overall Health - BL | 0.00 | 0.00 | 0.00 | 0.00 | 0.00 | 0.00 | 1.00 |
| Social Function - YR7 | Social Function - BL | 0.00 | 0.00 | 0.00 | 0.00 | 0.00 | 0.00 | 1.00 |
| Overall Health - BL | Physical Limit - YR4 | 0.00 | 0.00 | 0.00 | 0.00 | 0.00 | 0.00 | 1.00 |
| Physical Limit - BL | Quality of Life - BL | 0.00 | 0.00 | 0.00 | 0.00 | 0.00 | 0.00 | 1.00 |
| Physical Limit - BL | Social Function - YR4 | 0.00 | 0.00 | 0.00 | 0.00 | 0.00 | 0.00 | 1.00 |
| Physical Compared - YR4 | Quality of Life - YR4 | 0.00 | 0.00 | 0.00 | 0.00 | 0.00 | 0.00 | 1.00 |

*Note*. Numbers represent probability (0=low, 1=high) of edge type in GFCI network.

BL = Baseline, YR = follow up years (1-7)

Edge types:

--> 1) V1 is a driver of V2; AND 2) V1 and V2 are not confounded; AND 3) V2 is not a driver of V1.

<-- 1) V2 is a driver of V1; AND 2) V1 and V2 are not confounded; AND 3) V1 is not a driver of V2.

o--> 1) V1 drives V2; OR 2) V1 and V2 are confounded; OR 3) both are true.

<--o 1) V2 drives V1; OR 2) V1 and V2 are confounded; OR 3) both are true.

o--o 1) V1 drives V2; OR 2) V2 drives V1; OR 3) V1 and V2 are confounded; OR 4) Both 1) and 3) are true; OR 5) Both 2) and 3) are true. <--> 1) A latent variable drives V1 and V2; AND 2) V1 and V2 are not direct drivers of each other.
